# Sustained IFN signaling is associated with delayed development of SARS-CoV-2-specific immunity

**DOI:** 10.1101/2023.06.14.23290814

**Authors:** Elsa Brunet-Ratnasingham, Sacha Morin, Haley E. Randolph, Marjorie Labrecque, Justin Bélair, Raphaël Lima-Barbosa, Amélie Pagliuzza, Lorie Marchitto, Michael Hultström, Julia Niessl, Rose Cloutier, Alina M. Sreng Flores, Nathalie Brassard, Mehdi Benlarbi, Jérémie Prévost, Shilei Ding, Sai Priya Anand, Gérémy Sannier, Eric Bareke, Hugo Zeberg, Miklos Lipcsey, Robert Frithiof, Anders Larsson, Sirui Zhou, Tomoko Nakanishi, David Morrison, Dani Vezina, Catherine Bourassa, Gabrielle Gendron-Lepage, Halima Medjahed, Floriane Point, Jonathan Richard, Catherine Larochelle, Alexandre Prat, Nathalie Arbour, Madeleine Durand, J Brent Richards, Kevin Moon, Nicolas Chomont, Andrés Finzi, Martine Tétreault, Luis Barreiro, Guy Wolf, Daniel E. Kaufmann

## Abstract

Plasma RNAemia, delayed antibody responses and inflammation predict COVID-19 outcomes, but the mechanisms underlying these immunovirological patterns are poorly understood. We profile 782 longitudinal plasma samples from 318 hospitalized COVID-19 patients. Integrated analysis using k-means reveal four patient clusters in a discovery cohort: mechanically ventilated critically-ill cases are subdivided into good prognosis and high-fatality clusters (reproduced in a validation cohort), while non-critical survivors are delineated by high and low antibody responses. Only the high-fatality cluster is enriched for transcriptomic signatures associated with COVID-19 severity, and each cluster has distinct RBD-specific antibody elicitation kinetics. Both critical and non-critical clusters with delayed antibody responses exhibit sustained IFN signatures, which negatively correlate with contemporaneous RBD-specific IgG levels and absolute SARS-CoV-2-specific B and CD4^+^ T cell frequencies. These data suggest that the “Interferon paradox” previously described in murine LCMV models is operative in COVID-19, with excessive IFN signaling delaying development of adaptive virus-specific immunity.

## INTRODUCTION

Coronavirus disease 2019 (COVID-19) caused by severe acute respiratory syndrome coronavirus 2 (SARS-CoV-2) infection is a heterogeneous disease which ranges from asymptomatic infection to fatal outcome. Qualitative and kinetic differences in viral loads and immune responses have been associated with clinical severity: we (*1*) and others (*2–5*) have shown that SARS-CoV-2 plasma viral RNA (vRNA) levels predict fatality in COVID-19 patients. However, some patients succumbed to their infection in the absence of high plasma vRNA, while other individuals with high vRNA survived (*1*), indicating a role for additional factors.

High amounts of inflammatory cytokines have also been linked with fatal outcome (*6–8*). These cytokines are implicated in immunopathology since immunomodulatory treatments such as IL-6R antagonists (*9, 10*), systemic corticosteroids (*11*), and Janus kinase inhibitors (*4*) improve COVID-19 patients’ survival. Despite the well-established roles of interferon (IFN) pathways in priming of antiviral immunity and evidence that pre-existing defects of type I IFN responses are associated with adverse prognosis (*12, 13*), recombinant type I IFN therapy failed to improve COVID-19 outcomes (*14*) and may even be detrimental in severe disease (*15*). This apparent paradox is consistent with observations that sustained IFN levels impair lung healing (*16*), while their impact on adaptive immunity remains to be determined. In addition, delayed antibody responses (*17, 18*), possibly linked to disrupted coordination between virus-specific T and B cells (*19*), have been observed in patients with fatal disease.

Given the highly dynamic nature of COVID-19, binning patients based on clinical characteristics across the duration of their hospitalization can blur our understanding of the disease course. Although several studies have shown outcome associations with immunovirological feature(*7, 8, 20*), we still lack a global understanding of the immunovirological kinetics associated with disease heterogeneity. Endotypes, in which patients are grouped based on molecular rather than clinical characteristics, allow a more accurate identification of high-risk patient subsets (*21*). The interplay between these molecular signatures can be visualized and explored through dimensionality reduction. Potential of Heat-diffusion for Affinity-based Trajectory Embedding (PHATE) is a manifold learning algorithm which computes a nonlinear transformation of the data to represent the latent structure of a dataset in low dimensions (*22*). In parallel, a k-means algorithm can be used to group patients into defined clusters sharing similar features.

Using this type of integrative approach on cross-sectional measurements of plasma vRNA, Spike Receptor Binding Domain (RBD)-specific antibody responses, and plasma levels of cytokines and tissue damage markers, we identified four patient clusters corresponding to different systemic responses to acute SARS-CoV-2 infection. These endotypes closely associated with clinical severity. Longitudinal profiling and computational modeling of the antibody responses showed that delayed RBD-specific antibody response was a central feature in two clusters. Using whole-blood transcriptional profiling, we show that patients with this delay have sustained IFN signatures. These signatures were also negatively associated with SARS-CoV-2-specific CD4^+^ T cell and B cell responses, but not CD8^+^ T cell responses. These results highlight a role for excessive IFN signaling in disrupting adaptive humoral and cellular immune responses to a human viral infection.

## RESULTS

### Study design, patient characteristics and classification

We investigated prospectively enrolled hospitalized COVID-19 patients with symptomatic infection and a positive SARS-CoV-2 nasopharyngeal swab (NSW) PCR from two hospitals in Montreal, Québec, Canada (n=242, discovery cohort) and one in Uppsala, Sweden (n=76, validation cohort). Blood draws were serially done throughout their stay in-hospital at enrollment and at 2, 7, 14, and 30 days. Only acute samples, defined as those collected within 30 days of symptom onset (n=630 in the discovery cohort; 152 in the validation cohort), were considered. We previously observed that plasma vRNA, RBD-specific IgG antibodies, cytokines (TNFα, CXCL13, IL-6, IL-23, CXCL8, CCL2 and IL1Ra), and tissue damage markers [Receptor for Advanced Glycation Endproducts (RAGE), Angiopoietin-2 (Ang-2) and surfactant protein D (SP-D)] were associated to fatal outcome when measured 11 days after symptom onset (DSO11)(*1*), while associations with RBD-specific IgM and IgA levels did not reach statistical significance (*1*). For unsupervised data characterization, we considered these 14 measurements in cross-sectional samples taken at DSO11 (+/- 4 days, n = 242) (Fig 1A). We first visualized samples on a 2D scatter plot using the PHATE dimensionality reduction algorithm (*22*). In parallel, we performed a k-means clustering on the same data. We chose a cluster count of four, as running k-means with a higher number of clusters led to over-fragmentation and small clusters (n<20), preventing adequate subgroup characterization. The clustering resulted in two smaller clusters (1: n=38 and 2: n=49) and two larger ones (3: n=73 and 4: n=82), which strongly aligned with regions of the PHATE embedding (Fig 1B). No clinical or demographic data was used for computing the PHATE embedding and clustering.

**Figure 1.**
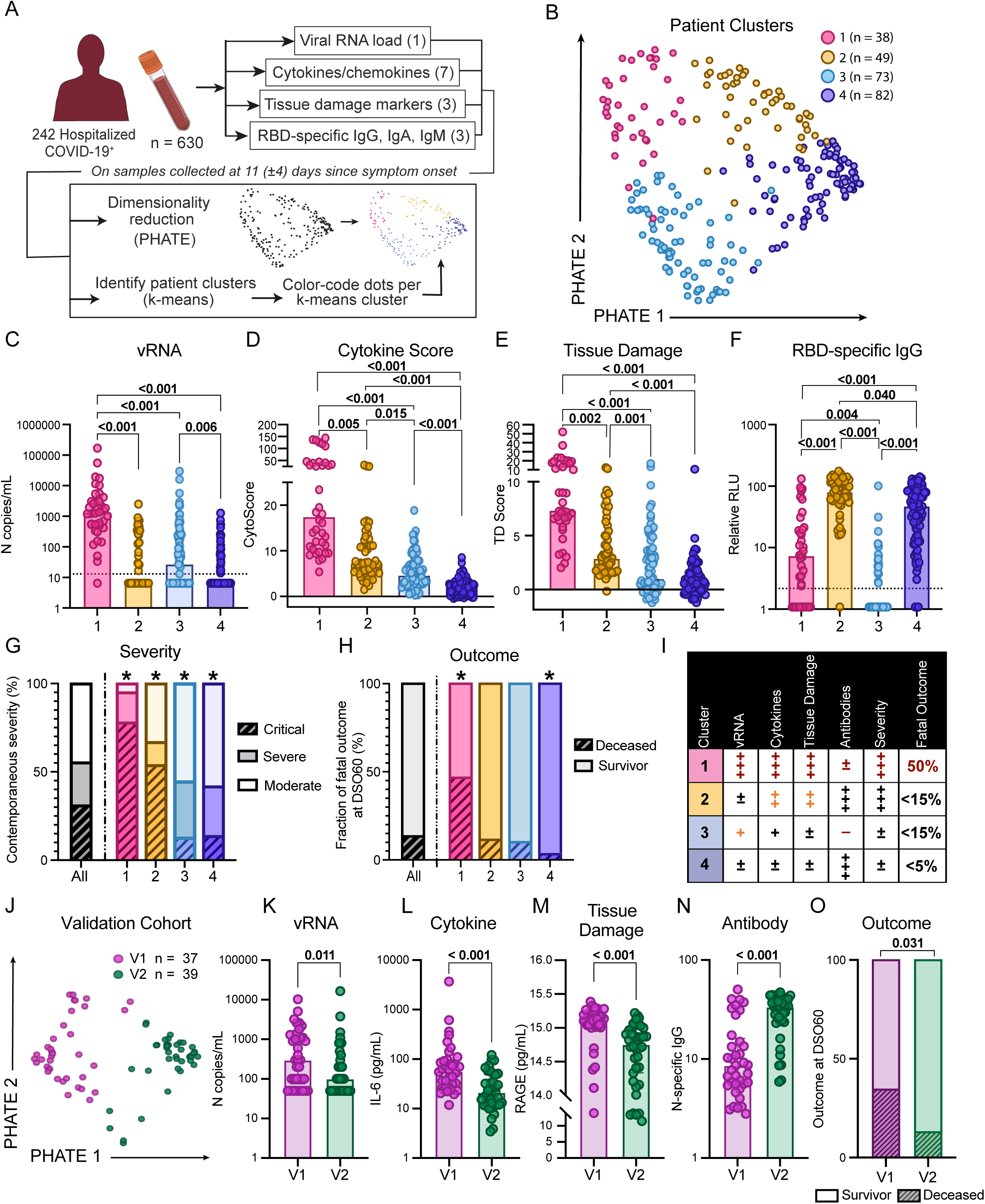
Hospitalized patients display four distinct endotypes of early plasma immunovirological profiles following SARS-COoV-2 infection. **A)** Study design. Serial blood samples were collected among hospitalized NSW PCR-confirmed COVID-19+ patients. Samples were assessed for plasma viral RNA, seven cytokines, three tissue damage markers, and three SARS-CoV-2 RBD-specific antibody isotypes. On samples collected 11 (+/- 4) days after symptom onset (DSO11), all 14 parameters were combined for visualization by PHATE and used to calculate patient clusters by k-means. Patient cluster was then used to color-code PHATE embedding. **B)** DSO11 samples identified four patient clusters across 242 hospitalized COVID-19 patients. **C-F)** At DSO11, plasma concentration across four patient clusters of **C)** viral RNA; **D)** Cytokine Score obtained from the linear combination of all seven cytokines; **E)** score of tissue damage obtained from the linear combination of all three markers of tissue damage, and **F)** SARS-CoV-2 RBD-specific IgG. **GH)** Percentage of the whole cohort or per patient cluster **G)** with critical (hashed), severe (saturated), or moderate (faint) disease; **H)** with fatal outcome (hashed). Chi^2^ compares the proportion of hashed groups in one cluster *versus* all others. **I)** Summary table of four patient clusters in the discovery cohort. **J)** Validation cohort of 76 hospitalized COVID-19 patients. SARS-CoV-2-specific IgG, IgM and IgA, vRNA and cytokines and tissue damage markers were measured at DSO11. PHATE embedding and k-means clustering were performed as for the discovery cohort. **K-O)** Comparison, between validation cluster (V)1 and V2, of plasma levels of K) SARS-CoV-2 vRNA; L) IL-6; M) RAGE or N) N-specific IgG. **O)** Outcome at DSO60 (hashed being fatal outcome). N discovery cohort: 1 = 38; 2 = 49; 3 = 73; 4 = 82 (242 in total). N validation cohort: V1 = 37; V2 = 39 (76 in total). CDEF) Kruskal-Wallis with Dunn’s multiple comparison tests. GH) Chi2 tests. KLMNO) Mann-Whitney tests.

### Hospitalized patients display four distinct endotypes of early plasma immunovirological profiles following SARS-CoV-2 infection

We examined how the parameters used to create the PHATE embedding differed between patient clusters at DSO11. Nearly all patients in cluster 1 had detectable vRNA and at higher amounts than the other clusters, with cluster 3 having the second-highest levels (Fig 1C). To integrate the overall quantities of cytokines, we created a cytokine score through the linear combination of the 7 cytokines surveyed (see Methods for details). This score followed a stepwise decrease between clusters 1, 2, 3, and 4 (Fig 1D). A second score created with the three markers of tissue damage also showed a stepwise decrease from cluster 1 to 4, although there were no differences between clusters 3 and 4 (Fig 1E). At this DSO11 timepoint, the RBD-specific IgG response was high in clusters 2 and 4, low in cluster 1, and undetectable in most subjects of cluster 3 (Fig 1F), with analogous patterns observed for RBD-specific IgM (Fig S1A) and IgA (Fig S1B).

To assess how these different immunovirological patterns associated with disease severity, we examined the contemporaneous patient status based on the level of respiratory support received (Moderate = no supplemental oxygen; Severe = oxygen on nasal cannula; Critical = non-invasive or invasive mechanical ventilation). Clusters 1 and 2 were enriched for critical patients, while clusters 3 and 4 mainly contained non-critical patients (Fig 1G, Fig S1C). Cluster 1 identified the most severe cases, as reflected by outcome: 50% of patients in cluster 1 died within 60 days of symptom onset, while fatal outcome was observed in a minority of the other three clusters (Fig 1H, Fig S1D). This was also reflected in the duration of hospitalization (S1E). Age (Table 1, Fig S1F) and sex (Fig S1G) did not significantly differ among the four clusters after correction for multiple comparisons, and similar distributions of ethnicities were observed as well (Fig S1H). Other demographic characteristics were largely similar between cohorts, except for the enrichment of pre-existing renal failure in cluster 1 (Table 1).

**Table 1.** Demographics and characteristics of hospital stay per patient cluster of the Discovery cohort. Values displayed are medians, with IQR in parentheses for continuous variables, or percentages for categorical variables. Percentages were rounded to the nearest unit. Statistical comparison across all four patient clusters, with Kruskal Wallis test for continuous variables, and *χ*^2^ test for categorical variables.

| Variable | Entries | All | 1 | 2 | 3 | 4 | Stats |
| --- | --- | --- | --- | --- | --- | --- | --- |
| n |  | 242 | 38 | 49 | 73 | 82 |  |
| Age | [median (IQR<sup>#</sup>)] | 66.4<br>(52.4 - 78.8) | 71.8<br>(64.4 - 78.8) | 66.9<br>(58.3 - 79.5) | 64.8<br>(54.1 - 82.0) | 57.6<br>(48.2 - 74.3) | **0.032** |
| Sex |  |  |  |  |  |  |  |
|  | Male | 141 (58%) | 25 (66%) | 27 (55%) | 38 (52%) | 51 (62%) | 0.43 |
|  | Female | 101 (42%) | 13 (34%) | 22 (45%) | 35 (48%) | 31 (38%) |  |
| Max respiratory Support throughout hospital stay |  |  |  |  |  |  |  |
|  | No O<sub>2</sub> | 86 (36%) | 5 (13%) | 10 (20%) | 36 (49%) | 35 (43%) | **< 0.001** |
|  | NC | 63 (26%) | 5 (13%) | 8 (16%) | 22 (30%) | 28 (34%) | **0.023** |
|  | NIV | 32 (13%) | 7 (18%) | 9 (18%) | 5 (7%) | 11 (13%) | 0.2 |
|  | ETI | 59 (24%) | 20 (53%) | 22 (45%) | 10 (14%) | 7 (9%) | **< 0.001** |
|  | ECMO | 2 (1%) | 1 (3%) | 0 (9%) | 0 (0%) | 1 (1%) | 0.44 |
| Days of hospitalization [median (IQR)] |  | 14 (8.0 - 27.0) | 24 (10.0 - 36.0) | 19 (12 - 41.5) | 14 (7.0 - 25.0) | 9 (5.3 - 15.8) | **< 0.001** |
| ICU admission |  | 89 (37%) | 27 (71%) | 29 (59%) | 15 (21%) | 18 (22%) | **< 0.001** |
| Days in ICU [median (IQR)] |  | 14 (5.0 - 31.0) | 24 (8.5 - 34.0) | 19 (7.0 - 35.0) | 14 (9.0 - 25.5) | 4.5 (3.3 - 6.8) | **< 0.001** |
| Metabolic risk factors |  |  |  |  |  |  |  |
|  | None | 75 (31%) | 10 (26%) | 14 (29%) | 24 (33%) | 27 (33%) | 0.85 |
|  | One or more | 167 (69%) | 28 (74%) | 35 (71%) | 49 (67%) | 55 (67%) |  |
|  | Obese | 33 (14%) | 5 (13%) | 12 (24%) | 10 (14%) | 6 (7%) | 0.053 |
|  | Hypertension | 139 (57%) | 26 (68%) | 30 (61%) | 41 (56%) | 42 (51%) | 0.32 |
|  | Dyslipidemia | 57 (24%) | 11 (29%) | 16 (33%) | 14 (19%) | 16 (20%) | 0.22 |
|  | Diabetes | 86 (36%) | 15 (39%) | 19 (39%) | 27 (37%) | 25 (30%) | 0.69 |
| Chronic diseases |  |  |  |  |  |  |  |
|  | None | 99 (41%) | 9 (24%) | 19 (39%) | 32 (44%) | 39 (48%) | 0.089 |
|  | One or more | 143 (59%) | 29 (76%) | 30 (61%) | 41 (56%) | 43 (52%) |  |
|  | Chronic Kidney Disease | 41 (17%) | 13 (34%) | 9 (18%) | 10 (14%) | 9 (11%) | **0.013** |
|  | Heart Failure | 51 (21%) | 11 (29%) | 11 (22%) | 15 (21%) | 14 (17%) | 0.52 |
|  | Respiratory Disease | 42 (17%) | 12 (32%) | 8 (16%) | 9 (12%) | 13 (16%) | 0.078 |
|  | Liver Disease | 21 (9%) | 7 (18%) | 5 (10%) | 6 (8%) | 3 (4%) | 0.63 |
|  | Immunosuppressed | 20 (8%) | 4 (11%) | 6 (12%) | 6 (8%) | 4 (5%) | 0.47 |
|  | Malignancy | 35 (14%) | 5 (13%) | 7 (14%) | 10 (14%) | 13 (16%) | 0.98 |
|  | HIV | 5 (2%) | 2 (5%) | 1 (2%) | 1 (1%) | 1 (1%) | 0.5 |
|  | Neurological disorder | 30 (12%) | 3 (8%) | 8 (16%) | 8 (11%) | 11 (13%) | 0.65 |
| Risk factors (Metabolic + Chronic) |  |  |  |  |  |  |  |
|  | None | 40 (17%) | 4 (11%) | 9 (18%) | 13 (18%) | 14 (17%) | 0.75 |
|  | One or more | 202 (83%) | 34 (89%) | 40 (83%) | 60 (82%) | 68 (83%) |  |
| Outcome |  |  |  |  |  |  |  |
|  | Fatality DSO60 | 34 (14%) | 18 (47%) | 6 (12%) | 7 (10%) | 3 (4%) | **< 0.001** |

Our analytical approach therefore identified four immunovirological endotypes in SARS-CoV-2 infection at DSO11 (Fig 1I) that not only aligned with contemporaneous disease severity but also delineated probability of survival among critical cases.

### Replication of the high fatality cluster 1 in an external validation cohort

The distinct validation cohort, recruited in a Swedish hospital, differed to clusters 1 and 2 of the discovery cohort for age, sex, and prevalence of some pre-existing conditions (Table 2). The validation cohort had a greater incidence of mechanical respiratory support, in line with recruitment of this cohort exclusively from the ICU (Table 2). Despite these differences, the incidence of fatal outcome was similar. The PHATE and k-mean analyses were performed using a subset of measurements common between both the discovery and validation cohorts, including the 3 antibody measurements, plasma vRNA, 5 of 7 cytokines, and 1 of 3 tissue damage markers (see Methods for details). PHATE produced two natural clusters that again aligned with k-means analysis (Clusters V1 and V2) (Fig 1J). The two clusters recapitulated the immunovirological patterns identified in clusters 1 and 2 in the discovery set. Cluster V1 showed higher viral load (Fig 1K), inflammation (Fig 1L), and tissue damage (Fig 1M). The differences in vRNA and tissue damage markers were less pronounced, likely due to differences in measurement methods and the use of only a subset of analytes compared to the discovery cohort. Cluster V1 also had lower N-specific antibodies at DSO11 (Fig 1N) than cluster V2. Nonetheless, cluster V1 was strongly enriched in fatal outcome (Fig 1O). The strong reproducibility between both cohorts, even with the use of a subset of the original set of analytes and differences in the SARS-CoV-2 target of the antibodies measured, validates the use of the PHATE/k-means analysis of early plasma profile to classify patient heterogeneity.

**Table 2.** Comparison of demographics and characteristics of hospital stay between critical clusters 1 and 2 of Discovery cohort with the validation cohort. Values displayed are medians, with standard deviation in parentheses for continuous variables, or percentages for categorical variables. Percentages were rounded to the nearest unit. Unpaired *t* test for continuous variables, and *χ*^2^ test for categorical variables.

| Variables | Entries | Cluster 1 + 2 -<br>Discovery | Validation (entire<br>cohort) | p |
| --- | --- | --- | --- | --- |
|  | N | 87 | 76 |  |
|  | Age (mean (SD)) | 68.8 (13.72) | 59.42 (13.75) | **< 0.001** |
|  | Sex (male (%)) | 52 (59.8%) | 60 (78.9) | **0.0084** |
| Max respiratory support throughout hospital stay |  |  |  |  |
|  | No O<sub>2</sub> | 15 (17.2%) | 0 (0%) | **< 0.001** |
|  | NC/HFNC | 13 (14.9%) | 0 (0%) | **< 0.001** |
|  | NIV | 16 (18.4%) | 25 (32.9%) | **0.033** |
|  | ETI | 42 (48.3%) | 49 (65.3%) | **0.038** |
|  | ECMO | 1 (1.2%) | 2 (2.6%) | 0.48 |
|  | ICU admission | 56 (64.4%) | 76 (100%) | **< 0.001** |
|  | Days in ICU (mean (SD)) | 20.5 (20.55) | 14.54 (10.04) | **0.023** |
| Metabolic risk factors |  |  |  |  |
|  | Any metabolic risk factor | 63 (72.4%) | 50 (75.8%) | 0.36 |
|  | Obese (%) | 17 (19.5%) | 27 (40.9%) | **0.022** |
|  | Hypertension (%) | 56 (64.3%) | 40 (52.6%) | 0.13 |
|  | Diabetes (%) | 34 (39.1%) | 21 (27.6%) | 0.12 |
| Chronic diseases |  |  |  |  |
|  | One or more (%) | 59 (67.8%) | 38 (50.7%) | 0.12 |
|  | Chronic Kidney Disease (%) | 22 (25.3%) | 17 (22.4%) | 0.66 |
|  | Heart Failure (%) | 22 (25.3%) | 3 (3.9%) | **< 0.001** |
|  | Respiratory Disease (%) | 20 (23.0%) | 19 (25.0%) | 0.76 |
|  | Liver Disease (%) | 12 (13.8%) | 0 (0%) | **< 0.001** |
|  | Immunosuppressed (%) | 10 (11.5%) | 10 (13.3%) | 0.75 |
|  | Malignancy (%) | 12 (13.8%) | 3 (3.9%) | **0.03** |
|  | HIV (%) | 3 (3.4%) | 0 (0%) | 0.10 |
|  | Neurological disorder (%) | 11 (12.6%) | 4 (5.3%) | 0.10 |
| Risk factors metabolic or chronic |  |  |  |  |
|  | One or more (%) | 74 (85.1%) | 58 (89.2%) | 0.16 |
| Outcome |  |  |  |  |
|  | Fatality DSO60 (%) | 24 (27.8%) | 18 (23.7%) | 0.57 |

### Delayed antibody kinetics is associated with protracted plasma vRNA over a wide range of disease severity

Given the differences in antibody levels among the discovery clusters at DSO11, we examined whether these resulted from either a delay or an inability in generating anti-RBD antibodies. We compared antibody levels at a later timepoint (DSO20 +/- 4 days) and saw no significant differences between patient clusters (Fig S2A), indicating a late, but ultimately comparable, response. To compute these differences in antibody kinetics, we combined the RBD-specific antibody measurements of all samples within a cluster and modeled a logistic curve on the quantity of antibodies per patient cluster per day (Fig 2A). Statistical comparison using bootstrap analysis (see Methods for details) revealed that patient clusters had significantly distinct timings in the generation of RBD-specific IgG responses. Both clusters 2 and 4 reached 50% of maximum RBD-specific IgG amount (DSO_50%_) before DSO11 (Fig 2A), which is why they had already high levels of antibodies at DSO11. Conversely, cluster 1 reached DSO_50%_ around DSO13, and cluster 3 was latest, with a DSO_50%_ at 17 days. This delayed antibody response was also observed in the validation cohort (Fig S2BC). We also measured the evolution of RBD-specific IgM (Fig S2D) and IgA (Fig S2E) and observed delayed responses in Clusters 1 and 3 that were similar to those observed for IgG. Thus, this delayed kinetics cannot be selectively attributed to impaired class switching.

**Figure 2.**
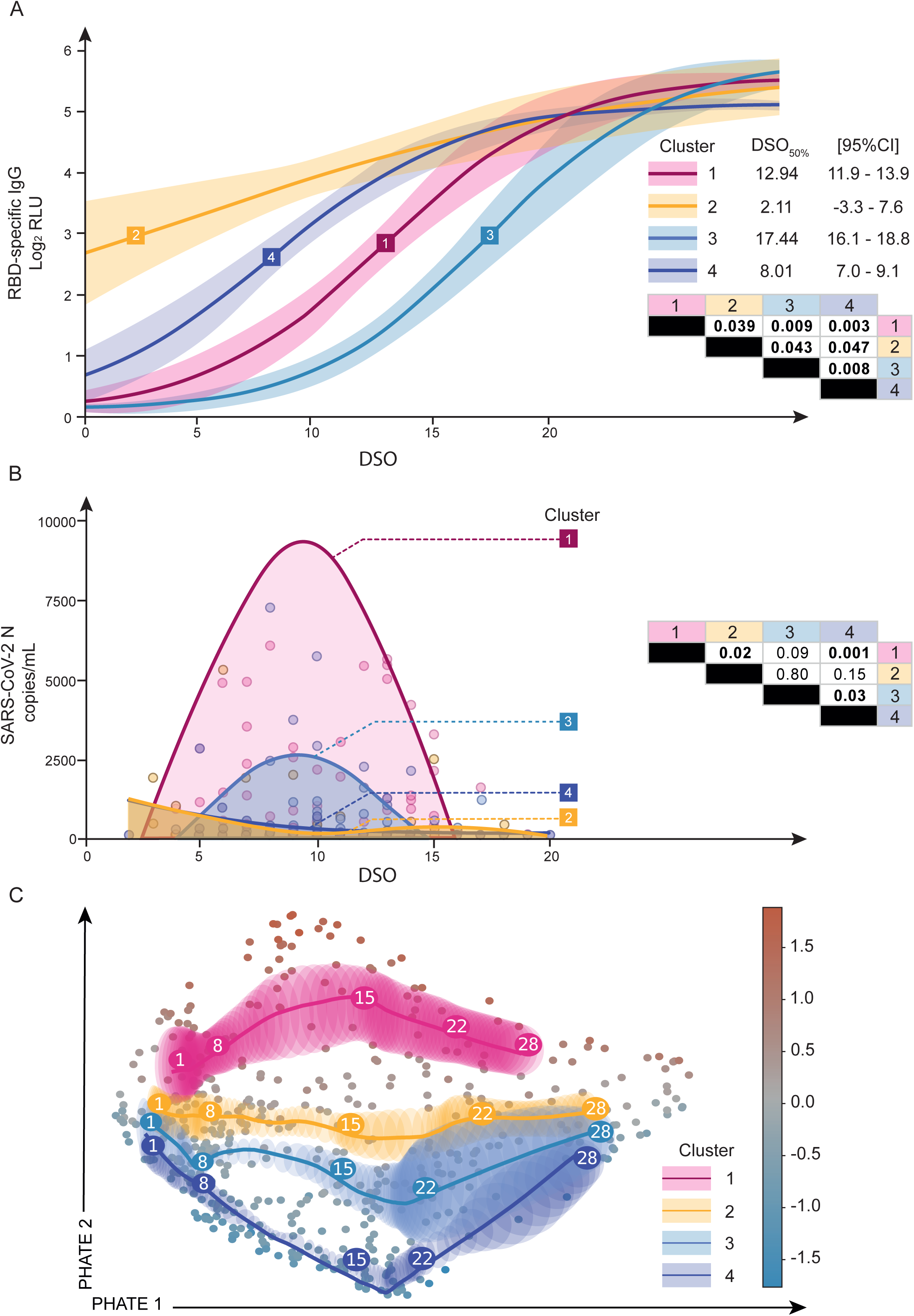
Delayed antibody kinetics is associated with protracted plasma vRNA over a wide range of disease severity. **A)** Sigmoidal curve fitted to the average per day per patient cluster of RBD-specific IgG responses. Squares represent coordinates where 50% of max IgG level is reached per cluster (DSO_50%_). Extrapolated DSO_50%_ value and 95% confidence intervals (CI) are on the right of the graph. P values were calculated using bootstrap comparison of DSO_50%_ in the table on the bottom right. **B)** Model of plasma vRNA detection, fitted to the average per day per patient cluster, among viremic patients only. Bootstrap on the area under the curve (AUC) was used to compare clusters, with p values provided in the table on the right of the graph. Faded dots represent raw data points per DSO. **C)** Average trajectory per color-coded patient cluster when the PHATE embedding was performed using cytokines (7), tissue damage markers (3) and DSO across all acute samples. Numbers in large circles represent the day of symptom onset at that coordinate. Shaded area represents confidence interval. Smaller circles in background are datapoints, color-coded by average analytes expression. AC) N = 630 data points. B) 224 datapoints (only RNAemia+ subjects were considered). AB) Two-stage bootstrap, with 1000 simulations. Pairwise comparison between all four clusters. C) Bootstrapping at the patient level was used to visualize the confidence ellipses representing 3 standard deviations around the average. See material and methods for details.

We next investigated whether there were also differences in plasma viral RNAemia throughout infection (herein referred to as vRNA exposure). 97% of cluster 1’s patients had detectable plasma vRNA at least once during their hospital stay, compared to 65% of cluster 2 patients, 55% of cluster 3, and 41% of cluster 4 (Fig S2F). We fitted a 4-knot spline curve onto the average vRNA per day per cluster and calculated the area under the curve (AUC) per clusters, as a metric for overall exposure (Fig 2B). Cluster 1’s vRNA AUC was significantly greater than that of clusters 2 and 4. Cluster 3 also had significantly greater vRNA exposure compared to cluster 4. Taken together, these results indicate that, among patients with detectable plasma vRNA, those with delayed antibody generation had greater overall exposure to plasma vRNA compared to their severity-matched counterparts.

To better understand how the plasma cytokine and tissue damage profiles evolved, we created a PHATE embedding using the 10 cytokine and tissue damage variables, with days since symptom onset (upweighted, methods for details) for all available data points within DSO28 of the discovery cohort (n = 242 subjects, 630 data points, Fig 2C). Marker color (gradient bar, right) reflects the average cytokines and tissue damage markers concentration of a given sample, unveiling a gradient from the low-concentration region (bottom) to a high concentration one (top). The average trajectories per cluster were plotted atop the embedding (see methods for details). They differed the most in the DSO8-15 interval, with cluster 1 exhibiting the highest cytokine levels and cluster 4 the lowest. We observed convergence of clusters 2,3 and 4 to a common region of the embedding by DSO28, consistent with transition to convalescence. Cluster 1 stood out as maintaining a high concentration of cytokines and tissue damage markers throughout the considered time period, in line with that cluster’s greater severity and higher ongoing inflammatory profile compared to the other three clusters.

Therefore longitudinal plasma profiling of hospitalized COVID-19 patients revealed that a delayed generation of RBD-specific antibodies coincided with greater viral exposure throughout the acute phase, suggesting that this delayed antibody response is important to COVID-19 pathogenesis. High and sustained levels of cytokine and tissue damage markers were hallmarks of critical disease.

### Fatal outcome among cluster 1 defined by specific transcriptomic signatures

To understand the molecular features behind the patient endotypes, we performed bulk RNA sequencing on 369 whole blood samples collected within 30 days of symptom onset, 174 of which were collected in the DSO11 timeframe (Table S1). Principal component analysis (PCA) on significant differentially expressed genes (DEG – False Discovery Rate (FDR) < 0.01, n = 3 271, Table S2) between all pairwise comparisons of the four clusters’ DSO11 samples revealed segregation of low (1 and 3) and high (2 and 4) antibody clusters along PC1 (Fig 3A).

**Figure 3.**
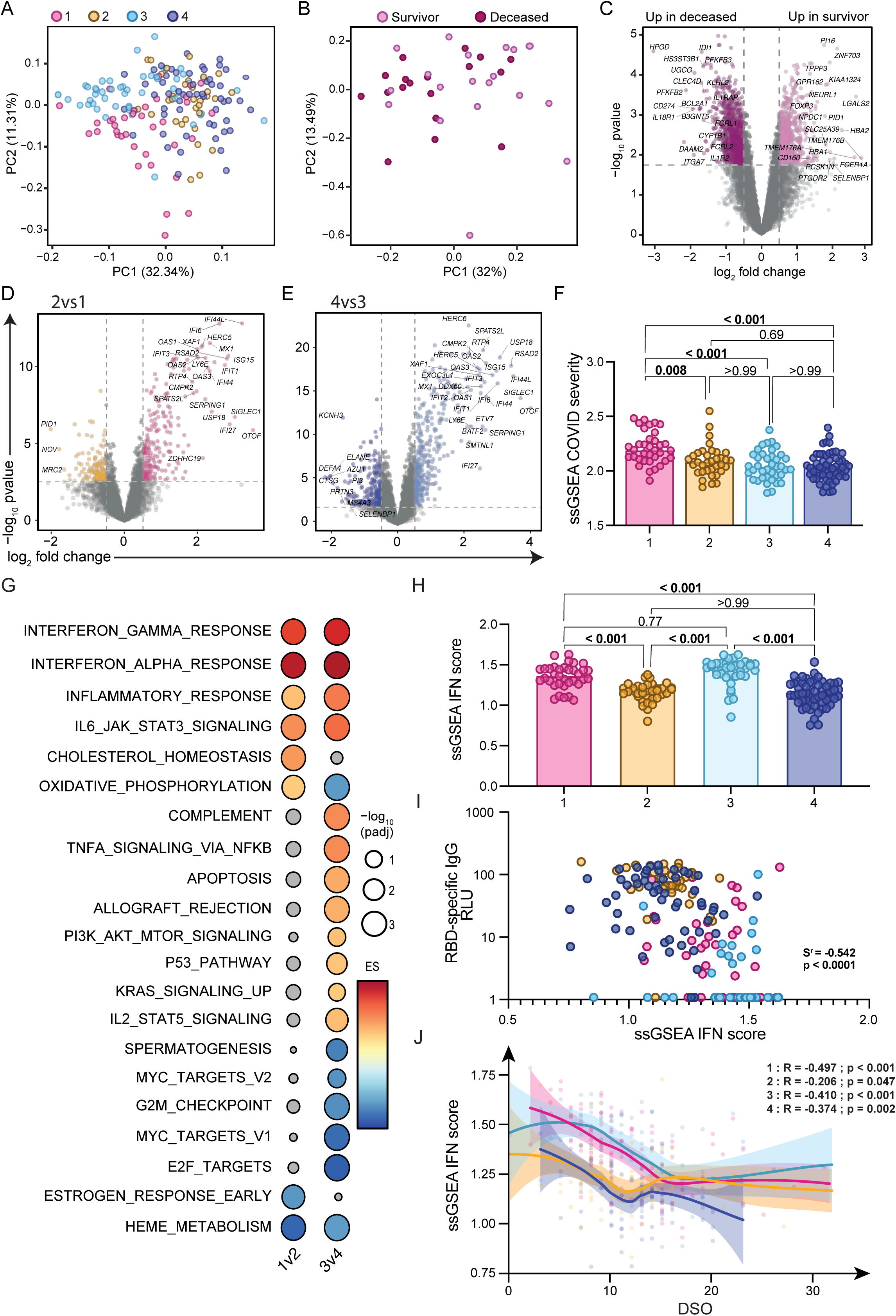
Patients with delayed SARS-CoV-2-specific antibody responses display sustained IFN signaling. **A)** Principal component analysis (PCA) based on significant DEG (FDR < 0.01, n = 1 346 genes) from all pairwise comparisons across the 4 patient clusters. Each dot represents a separate patient, sampled at DSO11, and color-coded to their respective cluster. **B)** PCA on whole transcriptome (n= 10 236 genes) of patients in cluster 1 only at DSO11, color coded by survival or fatal outcome at DSO60. **C**) Volcano plot of differentially expressed genes (DEG) based on outcome, with significant genes color-coded (FDR < 0.05; |logFC| > 0.5): mauve dots represent genes increased in fatal outcome, and pink, genes increased in survivors. Relevant genes are tagged. **DE**) Volcano plots of contrasts D) 1 vs 2 or E) 3 vs 4, with significant genes (FDR < 0.05; |logFC| > 0.5) color-coded and relevant genes tagged. **F**) Single sample (ss)GSEA of published COVID-19 severity score (*23*) across patient clusters. **G**) GSEA using Hallmark dataset on t-statistics from aforementioned contrasts. Red dots are pathways enriched in the low antibody clusters 1 and 3 compared to 2 and 4, respectively, while blue dots are pathways enriched in high antibody clusters. Significant hits are colored. Size of the circle is representative of significance of enrichment. **H**) ssGSEA IFN score calculated from the combination of the “interferon gamma response” and “interferon alpha response” Hallmark gene sets across patient clusters. **I**) Correlation between IFN score and contemporaneous RBD-specific IgG levels at DSO11. **J**) ssGSEA IFN score over time (DSO < 40) per patient cluster, with confidence intervals shaded. R and p values of each cluster are annotated at the bottom of the figure. N: 1 = 100; 2 = 93; 3 = 86, 4 = 98 (377 in total). EH) Kruskal-Wallis with Dunn’s multiple comparison tests. IJ) Spearman correlations.

PHATE embedding of cluster 1 showed the homogeneous distribution of fatal outcome (Fig S3A), in line with the absence of differences in plasma levels of vRNA (Fig S3B), cytokines (Fig S3C), tissue damage markers (Fig S3D), and in antibody responses (Fig S3E) between deceased and survivors of this cluster. PCA on the transcriptomic profiles of the survivor and deceased patients of cluster 1 showed substantial overlap (Fig 3B), although contrasting both outcomes revealed thousands of DEG (n = 1 537, FDR < 0.05, |logFC| > 0.05) (Fig 3C, Table S2). Our previously published COVID severity signature (*23*), aggregated into a single “score” per sample [single sample gene set enrichment analysis (GSEA) – ssCOVID] did not differ between outcome (Fig S3F). These results suggest that additional processes, rather than exacerbation of those associated with COVID-19 severity, contributed to fatalities in cluster 1. GSEA using Hallmark gene sets (*24*) revealed that, within cluster 1, deceased patients had increased signatures of TGFβ and mTOR signaling, while survivors had increased signatures of oxidative phosphorylation, MYC targets, coagulation, DNA repair and fatty acid metabolism (Fig S3G, Table S3). Thus, our results suggest that exacerbated inflammatory responses, when coupled with other mechanisms that include cell metabolism dysregulation, immunosuppression and fibrosis-related signaling (both roles of TGFβ), increase COIVD-19 fatality risk.

### Patients with delayed SARS-CoV-2-specific antibody responses display sustained IFN signaling

To uncover the molecular mechanisms underlying the delayed antibody response, we compared whole blood transcriptomes between clusters according to anti-RBD antibody responses at DSO11. To account for the impact of disease severity on transcriptomic profiles, we performed pairwise comparisons between low and high antibody patient clusters stratified by clinical status: we compared critical clusters 1 *versus* 2, and non-critical clusters 3 *versus* 4. Hundreds of genes were differently associated (FDR < 0.05, |logFC| > 0.5) with antibody status for both comparisons (clusters 1 vs 2 DEGs = 400; clusters 3 vs 4 n DEGs = 674, Fig 3DE, Table S2). The COVID severity score was increased in cluster 1 compared to all other clusters (Fig 3F). Various immune signatures were enriched among genes displaying higher expression in the low-antibody response clusters 1 and 3, most notably the IFN gamma response and IFN alpha response pathways (FDR < 2e-4, Fig 3G, Table S3). Compared to its high-antibody counterpart, cluster 3 had increased signatures of complement and TNFα signaling, and cluster 1 had increased oxidative phosphorylation (Fig 3G). Gene ontology enrichments on the Biological processes (*25*) similarly showed that patient clusters with delayed antibody responses were enriched for pathways related to IFN signaling and to the defense against invading pathogens (Fig S3H, Table S3), with cluster 3 further enriched for pathways related to regulation of cytokine production and TLR7 signaling (Fig S3I, Table S3). Overall, the transcriptomic profile of low-antibody clusters was characterized by heightened defense pathways against pathogens and type I and II IFN signatures.

To further investigate these IFN signatures, we next performed single sample GSEA combining all genes of both IFN gamma/alpha response pathways into a single score (ssIFN, Fig 3H). In line with the enrichment analyses, clusters 1 and 3 had significantly higher ssIFN scores than clusters 2 and 4, respectively. The ssIFN score showed a strong negative correlation with contemporaneous RBD-specific IgG levels (Fig 3I), and showed weaker positive correlations with contemporaneous plasma vRNA (Fig S3J), cytokine scores (Fig S3K), and tissue damage scores (Fig S3E). Thus, sustained IFN responses are associated with multiple immunopathological traits in COVID-19.

We next plotted the IFN score over days since symptom onset per patient cluster (Fig 3J). This analysis revealed that clusters 1 and 3 had high and sustained IFN for a longer period than the two other patient clusters, although all clusters converged around DSO15.

These results show that sustained upregulation of IFN pathways was associated with the delayed generation of SARS-CoV-2-specific antibody responses, this in patients exhibiting a wide range of disease severity.

### High IFN signaling is negatively associated with blood RBD-specific B cell and plasmablast frequencies

To investigate the cellular basis of poor antibody responses in the low-antibody patient clusters, we examined the blood SARS-CoV-2-specific B cell and plasmablast (PB) populations (Fig S4A). Staining peripheral blood mononuclear cells (PBMCs) with two fluorescently-labeled recombinant RBD probes identified RBD-specific B cells (Fig 4A) and PB (Fig S4B). While RBD-specific B cells were detectable in convalescent patients (Fig S4CD), RBD-specific PB were only detectable during acute infection (Fig S4EF), in line with the kinetics of circulating PB in COVID-19 (*26*). To account for lymphopenia in COVID-19 patients (*26*), we used contemporaneous clinical complete blood counts (CBC) to calculate the absolute frequencies of RBD-specific B cells and PB per mL of blood. Both populations correlated positively with each other (Fig 4B) and with RBD-specific IgG levels (Fig 4CD), consistent with their role in antibody production. B cell frequencies negatively correlated with the contemporaneous ssIFN score (Fig 4E), but no significant correlation was found for the PB (Fig 4F). Neither population correlated with the ssCOVID severity score (Fig S4GH). The cluster-level patterns were consistent with the strength of the IFN signatures (Fig 4G): cluster 3 had lower counts, and clusters 2 and 4 greater counts of SARS-CoV-2-specific B. For PB, differences did not reach statistical significance, as the spread within clusters was more pronounced (Fig 4H). There were no differences between acute infection clusters in isotype expression by RBD-specific B cells, which were mostly IgM+ and/or IgG+ (Fig S4I). This pattern differed from a separate cohort of convalescent outpatients (DSO > 100), in whom RBD-specific B cells were almost exclusively IgG+. RBD-specific PB displayed similar trends, albeit with a greater representation of IgG/IgA double-positive than their B cell counterparts (Fig S4J).

**Figure 4.**
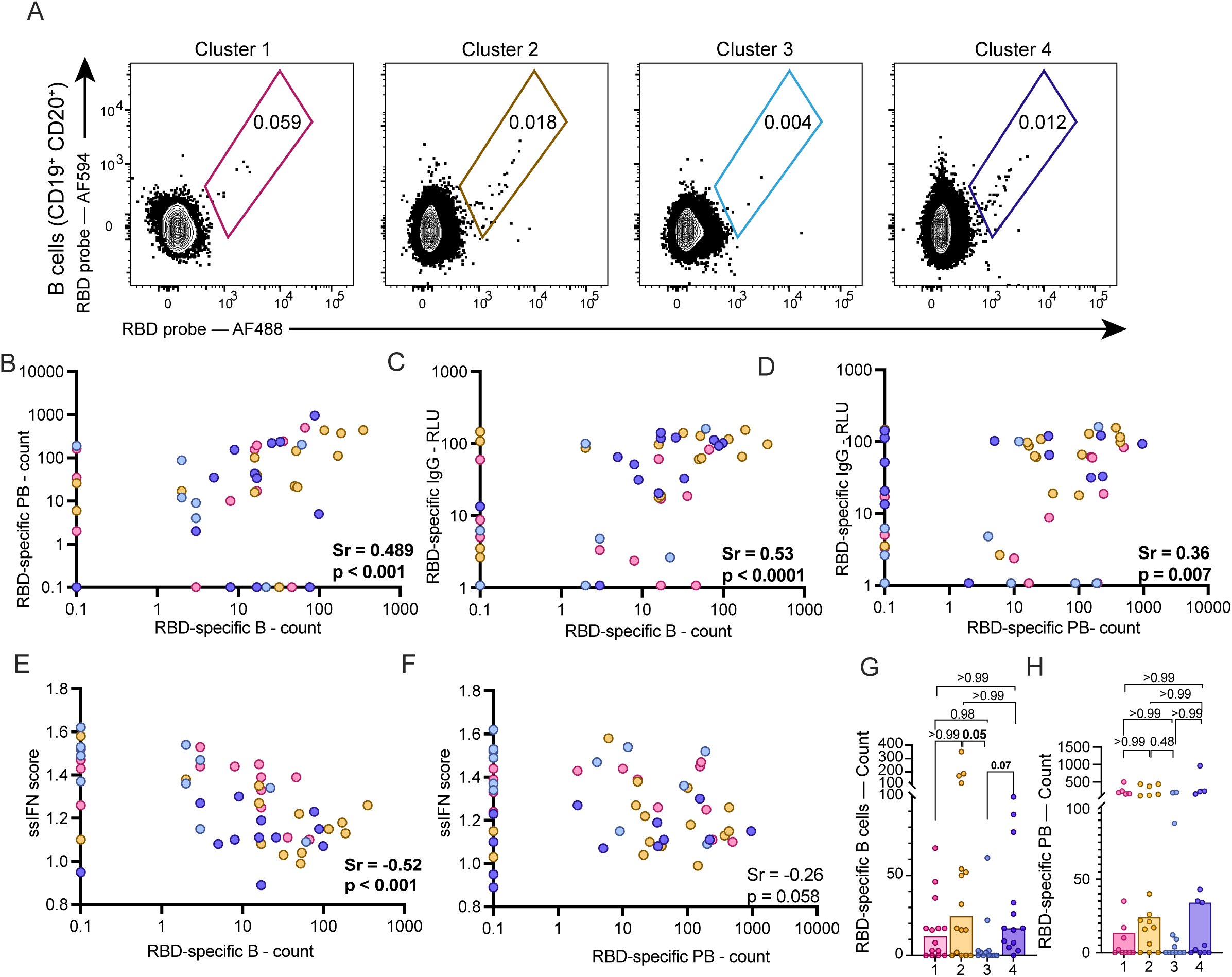
Elevated IFN signaling is negatively associated with RBD-specific B cell and plasmablast frequencies. **A)** Representative flow cytometry plots of RBD-specific B cells captured per patient cluster at DSO11. **B-F)** Correlation between B) absolute counts of RBD-specific B cells and RBD-specific PB; C) absolute counts of RBD-specific B cells and RBD-specific plasma IgG levels; D) absolute counts of RBD-specific PB cells and RBD-specific plasma IgG levels; E) absolute counts of RBD-specific B cells and ssGSEA IFN score; B) absolute counts of RBD-specific PB cells and ssGSEA IFN score. **GH)** Per patient cluster, absolute counts of RBD-specific G) B cells or H) PB. n for cluster 1 = 14; 2 = 16; 3 = 12; 4 = 13. BCDEF) Spearman correlations. GH) Kruskal-Wallis with Dunn’s multiple comparison tests. For patients with undetectable RBD-specific B and/or PB counts, they were assigned value 0.1.

Taken together, these results suggest that the sustained IFN signaling may hamper the generation of SARS-CoV-2-specific B cells, and consequently delay antibody responses in patients with various clinical presentations.

### High IFN signaling is negatively associated with Spike-specific CD4^+^ T cell responses

As persistent IFN signaling impairs adaptive virus-specific Thelper immunity in murine models (*27, 28*), we next examined the links between IFN transcriptional signatures and development of SARS-CoV-2-specific T cell responses. During acute SARS-CoV-2 infection, immunodominant peptides recognized by CD4^+^ T cells were mainly in the Spike-derived peptide pool, which also elicited CD8^+^ T cell responses in a majority of patients (Fig S5A-C). We measured the T cell responses against Spike using an activation-induced marker (AIM) assay we previously described (*29*). Spike-specific CD4^+^ T cells were detected by co-upregulation of CD69 with CD40L or OX40. We used a Boolean OR gating strategy to include overlapping populations (Fig 5A, Fig S5D). We again calculated absolute counts of Spike-specific CD4^+^ T cells by using CBC.

**Figure 5.**
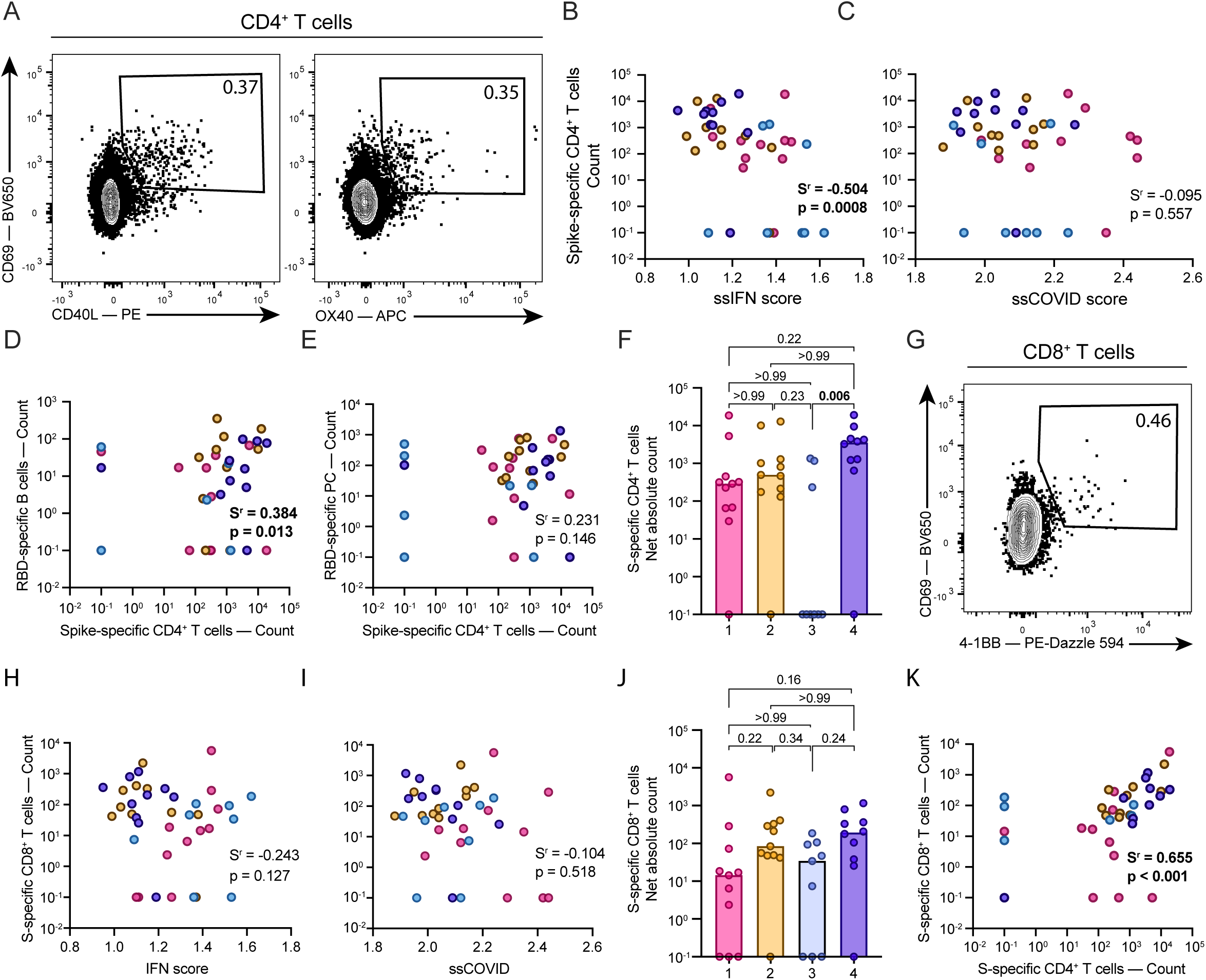
Elevated IFN signaling is negatively associated with Spike-specific CD4^+^ T cell responses. **A)** Representative flow cytometry gates used to detect Spike-specific CD4+ T cells following 15h peptide stimulation. Boolean OR gating strategy used. **B-E)** Correlations between absolute counts of Spike-specific CD4+ T cells with B) ssGSEA IFN score; C) ssGSEA COVID-19 severity score; D) absolute counts of RBD-specific B cells or E) absolute counts of RBD-specific plasma cells. **F)** Comparison of absolute counts of Spike-specific CD4^+^ T cells per patient cluster. **G)** Representative flow cytometry gate used to detect Spike-specific CD8_+_ T cells following 15hrs peptide stimulation. **H-I)** Correlations between absolute counts of Spike-specific CD8+ T cells with H) ssGSEA IFN score and I) ssGSEA COVID-19 severity score. **J)** Comparison of absolute counts of Spike-specific CD8+ T cells per patient cluster. **K)** Correlation between the absolute counts of both Spike-specific T cell populations. n for cluster 1 = 11; 2 = 11; 3 = 9; 4 = 10. BCDEHIK) Spearman correlation. FJ) Kruskal-Wallis with Dunn’s multiple comparison tests.

Spike-specific CD4^+^ T cells were detectable in most acute and all convalescent samples (Fig S5E). These frequencies correlated negatively with the ssIFN score (Fig 5B), suggesting a negative impact of IFN signaling on Thelper responses. There was no correlation between CD4^+^ T cell responses and ssCOVID severity scores (Fig 5C). Spike-specific CD4^+^ T cells counts positively correlated with RBD-specific B cells counts (Fig 5D), in line with the role of CD4^+^ T cell help in B cell immunity. No significant correlation was observed between Thelper responses and RBD-specific PB (Fig 5E). Spike-specific CD4^+^ T cell responses were detected in most patients in all clusters, except for cluster 3 (Fig 5F), underscoring a defect in adaptive T helper responses and paralleling the defects identified for RBD-specific B cells.

We measured Spike-specific CD8^+^ T cells by their co-upregulation of CD69 and 41BB (Fig 5G). Spike-specific CD8^+^ T cells were also detectable in most acute and all convalescent samples, although they were less frequent than their CD4^+^ T counterparts (Fig S5F). They correlated neither with ssIFN nor with ssCOVID scores (Fig 5HI), nor did they differ across clusters (Fig 5J). Despite the differential association of Spike-specific CD4^+^ and CD8^+^ T cell responses with ssIFN signatures, the two cell populations themselves correlated positively(Fig 5K), consistent with the role of CD4^+^ T cell help in primary CD8^+^ T cell responses.

Taken together, these results suggest that sustained IFN signaling negatively impacts SARS-CoV-2-specific CD4^+^ T cell responses, which in turn hamper the generation of SARS-CoV-2-specific B cells.

## DISCUSSION

Through a relatively simple immunovirological plasma profile 11 days after symptom onset (14 analytes: RNAemia, seven cytokines, three issue damage markers and three RBD-specific antibody isotypes), we identified COVID-19 patient endotypes with important differences not only in disease severity and outcome, but also in the quantity and timing of innate, antibody, and cellular responses to SARS-CoV-2. We also found that excessive IFN signaling likely contributes to differential kinetics of virus-specific antibody, B cell and CD4^+^ T cell responses. Early robust antibody and T cell immunity, coupled with a low inflammatory profile and low plasma viremia, is associated with moderate disease and good prognosis (cluster 4). When robust SAR-CoV-2-specific immune responses are maintained in the setting of a higher inflammatory profile, as typically observed in critical disease, the prognosis is still good (cluster 2). Patients with sustained IFN signaling also have delayed antibody and CD4^+^ T cell responses, which can be associated with a good prognosis if viremia and inflammation are low or moderate (cluster 3). However, sustained IFN signaling coupled with high viremia, exacerbated inflammatory profile and low SARS-COV-2-specific B cell and CD4^+^ T cell responses is associated with the distinctively high-fatality cluster 1.

Among critically ill patients, our approach delineated the subset of individuals at very high risk of fatality from those with unexpectedly good prognosis with greater accuracy compared to binning patients based on clinical severity, underlining the advantage of using immunovirological endotypes. The cluster-based method combined with model fitting and bootstrap pairwise comparisons made up for the sparsity of data points per patient, allowing for robust comparisons of trajectories in a time-dependent manner. The findings were highly reproducible in a separate cohort located in a different country, despite differences in the laboratory methods used. This robustness of the k-means algorithm, which builds on the high-dimensional relationships of features rather than absolute values, can be a major asset in multicentric translational biomedical research, where ensuring reproducibility of data can be complicated by the use of diverse technical platforms (e.g., institutional clinical lab instruments). For instance, despite measuring antibodies targetting different SARS-CoV-2 proteins (RBD for discovery; N for validation), both cohorts displayed low responses at DSO11 among the high fatality clusters. These results suggest an effect on the antibody response against SARS-CoV-2 as a whole, and underscore the value of a multiparametric approach in deciphering patient heterogeneity.

The RBD-specific antibody trajectory analyses revealed that the low antibody levels observed at DSO11 in clusters 1 and 3 were due to a delayed initiation of the antibody response to RBD, rather than an inability to do so. Indeed, the modeled curves converged prior to DSO30, consistent the robust SARS-CoV-2 specific antibody responses in convalescent individuals after critical disease. One unavoidable limitation here is possible survivorship bias, as we cannot determine if the critical cases who succumbed early in disease course had the potential to mount this response. These granular analyses also differentiate vRNA kinetics that do not merely align with clinical severity and were not identified by more traditional statistical tools (*1, 30*). While vRNA loads were highest in the high-fatality cluster 1, they were also sustained in the low-antibody, non-critical cluster 3, whereas both clusters that rapidly developed RBD-specific antibodies (2 and 4) readily achieved viral clearance. These results are consistent with the critical role of antibody responses in controlling viral replication. In outpatients, anti-Spike neutralizing monoclonal antibodies and antiviral drugs decrease the risk of disease progression only when given early after symptom onset (*31, 32*). There is also no or limited impact of monoclonal antibodies in people hospitalized for COVID-19 (*33*), except for a subset of those who have not yet seroconverted (*34*). These data indicate that once underway, the pathogenic inflammatory cascades, which diverge as early as DSO8, have limited sensitivity to these interventions.

Whole-blood transcriptional profiling provided important insight into the molecular features underpinning these endotypes. While the hierarchy of the COVID-19 severity signature we previously established (*23*) followed the clinical profile, pair-wise comparisons between the two critical clusters (1 vs 2) and between the non-critical clusters (3 vs 4) revealed differences in gene expression beyond disease severity. The pathways upregulated in low-antibody clusters 1 and 3, including IL-6-JAK-STAT3 signaling and other inflammatory pathways, are those for which blunting through therapies results in survival benefit (specifically by tocilizumab, sarilumab and baricitinib; and broadly by dexamethasone) (*4, 9, 11*), supporting a mechanistic explanation for these interventions. Signatures of complement activation and signaling through TLR7 and TNFα were only upregulated in cluster 3, suggesting that innate immunity may contribute to a moderate disease course despite sustained IFN signaling. Conversely, fatal cases of cluster 1 had further upregulation of IL-6-JAK-STAT3 signaling and inflammation, with exacerbated TNF signaling compared to cluster 1’s survivors. They also exhibited lower expression of MYC targets, which include many proliferation and anti-apoptotic pathways, along with depressed metabolic pathways. These findings suggest a disruption of key cellular processes in patients who subsequently succumb to their illness, which may help guide investigations of new therapeutic targets.

Because of potential mechanistic implications, a key novel finding of the transcriptional analyses is the differential kinetics of both type I and II IFN signatures among patient clusters. Although pronounced in cluster 1 fatalities, exacerbated IFN signatures did not merely coincide with disease severity, but rather with impaired generation of SARS-CoV-2-specific antibody, B cell and CD4^+^ T cell responses. These defects are probably causally linked, given the critical role of CD4^+^ T cell help for B cells (*35*). In contrast, we observed no significant associations with CD8^+^ T cell responses. These patterns suggest that protracted activation of IFN pathways can adversely affect some anti-SARS-CoV-2-specific responses, in addition to impairing repair mechanisms of damaged lung tissues (*36, 37*). Seminal studies in the murine lymphocytic choriomeningitis virus (LCMV) model support this hypothesis (*28, 38*) : while IFNs are critical in the early generation of antiviral responses, sustained type I IFN signaling in chronic Clone 13 infection was associated with poor antibody, B cell and CD4^+^ T cell responses. In this context, blockade of type I interferon signaling by an anti-IFNAR1 antibody decreased viral loads and improved immune responses. A similar benefit has recently been observed though the modulation of type I IFN in SARS-CoV-2 infection of rhesus macaques (*39*). This dual role of IFN, where timing, rather than quantity, is central to an appropriate adaptive response, is dubbed “The Interferon paradox” (*40*), and is also supported by data in SIV infection of non-human primates(*41*). In conjunction with data supporting a protective role of IFNs early in COVID-19 course (*12, 13*), our results suggest this paradox is operative in SARS-COV-2 infection in defined patient subgroups (even though, in contrast to LCMV Clone 13 and SIV, this major human viral disease is – with rare exceptions - an acute viral infection). This explains discrepancies around IFN in the literature. Individuals with inborn errors in type I IFN responses (*13*), genetic variants which lower IFN responsiveness (*42*) or pre-existing anti-IFN autoantibodies(*12*) are at greater risk of severe COVID-19 because they never have the initial IFN (key for reducing viral replication and the generation of an initial anti-viral response). However, the IFN signature should drop quickly, or it hampers the antiviral response and causes immunopathology. Patients are often put on mechanical ventilation around 9 days after symptom onset, so studies comparing critical and non-critical cases enrich in patients at those times, which is why high IFN signal was associated to severe disease in blood transcriptomics (*43*) and lung *in situ* (*44*) studies. It also explains the results from the clinical trial with IFN(*14*): it was administer too late to have any beneficial effect, and only the few people with genetic defects would have truly benefited from it.

In summary, we show that SARS-CoV-2-infected patients experiencing high, sustained IFN signaling have a delayed generation of Spike-specific CD4^+^ T cells and RBD-specific B cells. This directly links to a delay in the antibody response against the virus and, in patients also presenting increased inflammation, tissue damage, and plasma RNAemia, is associated with a highly fatal profile. Compared to mechanistic studies in mice, a weakness of the present observational study is the lack of direct manipulation of the type I IFN pathway. However, our results can have direct clinical relevance, and at least in part explain why clinical trials of recombinant IFN therapy have yielded disappointing results in COVID-19 (*45*). While our study lacked investigation of the lung compartment, others also support a pathophysiologic role of excessive type I IFN in this organ (*16, 36*). Hence, excessive type I IFN signaling is likely detrimental at multiple levels that involves adaptive immunity, and tissue repair. Whether targeted blockade of IFN pathways – rather than IFN supplementation – might be beneficial in specific subgroups of COVID-19 patients would require further investigation.

## MATERIAL AND METHODS

### Participants and samples

We investigated prospectively COVID-19 individuals hospitalized between April 2020 and August 2021 with symptomatic infection with a positive SARS-CoV-2 nasopharyngeal swab (NSW) reverse-transcription polymerase chain reaction (RT-PCR) who were admitted to the Centre Hospitalier de l’Université de Montréal (CHUM) or the Jewish General Hospital (JGH) and recruited into the Biobanque Québécoise de la COVID-19 (BQC19) (58) Blood draws were performed at baseline and, when consistent with patient care, at 2, 7, 14 and 30 days (±3 days) after enrollment. Exclusion criteria were breakthrough or reinfection, plasma transfer therapy (could change plasmatic profile), or vaccination prior to infection. The study was approved by the respective IRBs (multicentric protocol: MP-02-2020-8929). and written informed consent obtained from all participants or, when incapacitated, their legal guardian before enrollment and sample collection. Research adhered to the standards indicated by the Declaration of Helsinki. Blood draws were also performed on 50 asymptomatic, SARS-CoV-2 antibody negative uninfected controls (UC), early in the pandemic (spring 2020). COVID-19 hospitalized patients were stratified based on the severity of respiratory support at the DSO11 timepoint: critical patients required mechanical ventilation [noninvasive ventilation, endotracheal intubation, extracorporeal membrane oxygenation – (ECMO)], and non-critical patients, encompassing patients with moderate disease required no supplemental oxygen and patients with severe disease requiring oxygen supplementation by nasal cannula. Mortality was followed up to DSO60. Medical charts were reviewed by physicians and study coordinators for data collection on demographics, co-morbidities, risk factors, severity state, time of infection, etc. (see Table 1). Median age of the UC cohort was 37 years (range: 24-57), and 30 individuals were males (60%). Patient and sample identifiers were created in our research group and are not known to anyone outside our research group, as to protect the identity of the study subjects.

The validation cohort was recruited at Uppsala University Hospital in Sweden. The study was approved by the Swedish National Ethical Review Agency (Pronmed study; 2017-043, amended 2019-00169, 2020-01623, 2020-05730 and 2022-00526-01) and registered *a priori* at ClinicalTrials.gov (NCT03720860). Informed consent was obtained from the patient or next of kin if the patient was unable to give consent. The Declaration of Helsinki and subsequent revisions were followed. The study included 123 adult patients admitted to intensive care during the first wave of the pandemic between March 15^th^, 2020, and July 14^th^, 2020. All patients had confirmed SARS-CoV-2 by RT-PCR from NSW. Exclusion criteria were pregnancy, currently breastfeeding, and age under 18. A validation cohort of 76 patients was collected from the Pronmed study biobank with analyses that matched the parameters used for PHATE embedding in the discovery cohort.

### Measurements of plasma analytes

#### Quantification of plasma SARS-CoV2 RNA

For the discovery cohort, absolute copy numbers of SARS-CoV-2 RNA (N region) in plasma samples were measured by real-time PCR. Total RNA was extracted from 230 μL of plasma collected on acid citrate dextrose (ACD) tubes using the QIAamp Viral RNA Mini Kit (Qiagen Cat. No. 52906). Two master reaction mixes with specific primers and probes were prepared for quantification of N gene from SARS-CoV-2 and 18S (as a control for efficient extraction and amplification). A positive and no-template controls were included in all experiments. Purified RNA N transcripts (1328 bp) were quantified by Nanodrop, and the RNA copy numbers were calculated using the ENDMEMO online tool (see STAR methods for details).

For the external validation cohort, plasma viral RNA was determined by real-time RT-PCR recognizing the SARS-CoV-2 N-gene using the 2019-nCoV N1 reagent based on the Center for Disease Control (CDC) of the United States protocol as described previously (*3*).

#### Measurements of cytokines, chemokines and tissue damage markers

For the discovery cohort, duplicates of SARS-CoV-2-inactivated plasma samples were analyzed using a customized Human Luminex Discovery Assay (LXSAHM-26, R&D Systems). Datasets were acquired on two separate machines (BioPlex, MagPix), with 30 repeat samples performed on both. Linear regression was performed for each analyte and regressions used for batch correction of samples acquired on the BioPlex. As PHATE requires complete datasets, some analytes with low sensitivity that could not be corrected were excluded: CCL20, CCL3, CCL7, IFNα, GM-CSF, IL-10, IL-17A, IL-1b, IL-2, and IL-33. We retained all analytes significantly associated with fatal outcome (p<0.01) in our previous work (*1*): TNFα, CXCL13, IL-6, IL-23, CXCL8/IL-8, angiopoietin-2, RAGE, and Surfactant Protein D. For the validation cohort, plasma cytokines were measured using citrated plasma samples for 27 biomarkers with the Bio-plex assay using a Luminex MagPix instrument (Bio-Rad Laboratories AB, Sundbyberg, Sweden) as described previously (*6*). Of these 27, only the analytes in common with those measured in the discovery cohort were retained for the clustering and the PHATE embedding: TNFα, IL-6, CXCL8, IL1Ra, and CCL2. Plasma RAGE was also measured (and included as input to k-means and PHATE) using the Proximity extension assay (PEA) at the Clinical Biomarkers Facility (SciLifeLab, Uppsala, Sweden) using the Cardiovascular panel from OLINK Proteomics® (Uppsala, Sweden).

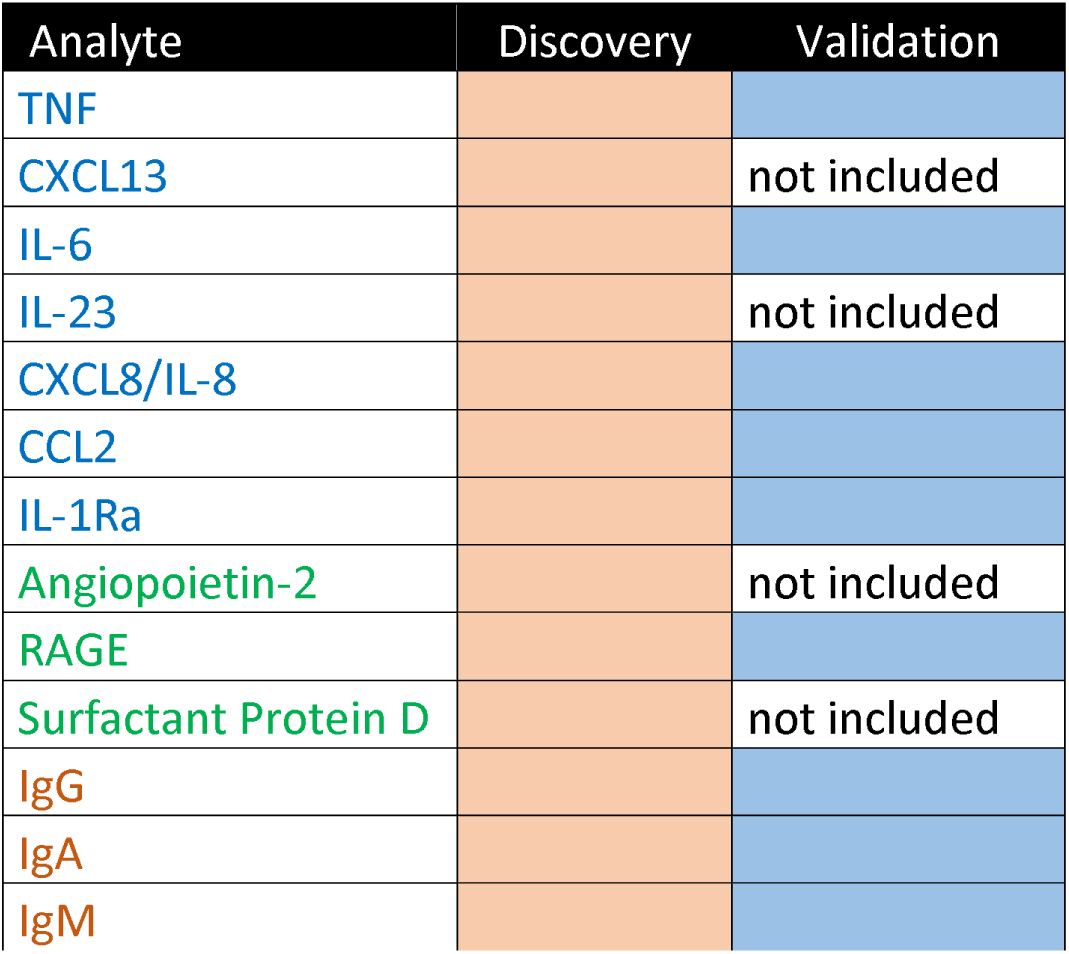
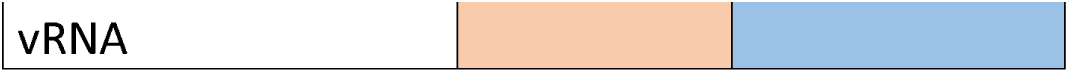

| Analyte | Discovery | Validation |
| --- | --- | --- |
| TNF |  |  |
| CXCL13 |  | not included |
| IL-6 |  |  |
| IL-23 |  | not included |
| CXCL8/IL-8 |  |  |
| CCL2 |  |  |
| IL-1Ra |  |  |
| Angiopoietin-2 |  | not included |
| RAGE |  |  |
| Surfactant Protein D |  | not included |
| IgG |  |  |
| IgA |  |  |
| IgM |  |  |

#### Antibody measurements

In the discovery cohort, RBD-specific IgG, IgM and IgA were quantified using an in-house SARS-CoV-2 RBD ELISA assay, as described elsewhere (*46*). Plasma from pre-pandemic uninfected donors were used as negative controls to calculate the seropositivity threshold in the ELISA assay. The monoclonal antibody CR3022 (*47*) was used as a positive control.. The seropositivity threshold was established using the following formula: mean of all COVID-19 negative plasma + (3 standard deviation of the mean of all COVID-19 negative plasma).

In the validation cohort, nucleocapsid-specific antibody levels at DSO11 were measured using the NovaLisa® SARS-CoV-2 IgA, IgM, and IgG kits according to manufacturer’s instructions (COVA0940, COVM0940, COVG0940, Novatec Immundiagnostica, Dietzenbach, Germany), and at DSO 20 using by FluoroEnzymeImmunoassay (FEIA), Phadia AB, Uppsala, Sweden as described previously (*17*).

### Data dimensionality reduction and clustering

#### PHATE

Dimensionality reduction is a necessary step to visualize and explore high dimensional datasets. While PCA(*48*) is commonly used, the resulting components are restricted to a linear projection of the input data, thereby limiting the expressiveness of the resulting visualizations. Recent advances in dimensionality reduction techniques instead favor so-called manifold learning algorithms, such as PHATE (Potential of Heat-diffusion for Affinity-based Trajectory Embedding), which can compute a nonlinear transformation of the data to effectively represent the latent structure of a dataset in low dimensions (*22*). PHATE begins by computing a sample-sample affinity graph, i.e., a graph connecting pairs of similar samples to form “neighborhoods”. The graph can then be leveraged to compute transition probabilities - the probability of a sample “jumping” to one of its’ graph neighbors in a random walk. By iteratively repeating this operation in a process known as diffusion, transition probabilities can be derived for any pair of samples in the dataset and therefore represent a useful notion of pairwise similarity, with probability transitions being high between close samples in terms of the diffusion geometry. Armed with these similarities, PHATE computes a two-dimensional embedding where the Euclidean distance reflects the dataset’s intrinsic structure as captured by diffusion, thus enabling the interpretation and analysis of high dimensional data Data samples are subsequently embedded in a low dimensional space (usually, 2 dimensions for visualization on scatter plots) by preserving both the local and long-range pairwise similarities, meaning the distance between “neighborhoods” in the embedding are meaningful. Intuitively, this can be thought as « unfolding » the sample-sample graph in low dimensions while preserving the graph’s intrinsic structure, as captured by diffusion affinities.

In practice, only explanatory variables (in our case, plasma concentrations) are used as input to PHATE and the structure of the embedding is therefore entirely unsupervised. PHATE then generates informative low-dimensional representations of the data and is known to preserve substructures of interest —such as clusters—while being robust to noise and non-uniform sampling of the underlying data manifold. Any variable of interest — such as patient outcome or clinical data — can be used for coloring. Particularly, explanatory variables can be used as color gradients to observe how they are distributed on the visualization (e.g., to identify high antibody and low antibody neighborhoods).

#### DSO11 PHATE embeddings

We computed PHATE visualizations of cross-sectional samples taken 11 days after symptom onset (DSO11 +/- 4 days) in the discovery cohort (n = 242, 14 measurements) and the validation cohort (n = 76, 10 measurements). Samples with missing measurements were removed, and if a given patient had more than one sample in the considered time period, only the one closest to DSO11 was included. Each 2D marker in the resulting scatter plots therefore summarizes the plasma profile of a single patient.

We use standard scaled (mean 0, variance 1) log concentrations of each sample as input to the PHATE Python package (v1.0.9) with a *knn* parameter of 10 for the discovery cohort and of 5 for the validation one. Both cohorts use a diffusion time *t* parameter of 50.

#### DSO11 K-means clustering

K-means (*49*) clustering aims to partition samples into different clusters with high intra-cluster similarity. The identified clusters can then serve as a basis for comparing typical groups or sample profiles. The method represents clusters as centroids (cluster centers), and iteratively refines them by alternating two steps: 1) assign samples to the closest centroids, and 2) replace centroids with the per-cluster sample means based on current assignments.

We clustered the same samples as the ones used for the DSO11 PHATE embeddings, using the k-means implementation of the scikit-learn(*50*) Python package (v1.0.2). We used 4 clusters for the discovery cohort and 2 clusters for the validation cohort due to the smaller number of samples. Data preprocessing was identical to the process used for PHATE.

#### MELD

Of particular interest to visualize binary outcome variables is the MELD algorithm(*51*), which performs a low-pass filtering of binary variables over the sample-sample graph to make them “smoother” over neighborhoods. The resulting values are used to compute relative likelihoods, thereby indicating if some groups of similar samples are enriched or depleted in a specific condition. In practice, this can be used to turn a binary variable into a continuous gradient which can be visualized on top of a PHATE embedding. We used MELD (v1.0.0) to obtain smooth visualizations of critical severity and fatal outcome in the DSO11 discover cohort.

#### Longitudinal embedding

We visualized the evolution of cytokine and tissue damage profiles in the discovery cohort using a second PHATE longitudinal embedding. The time horizon was increased to include all samples from DSO0 to DSO28. Only samples from patients selected for the DSO11 analysis were considered to better understand the progression of the identified DSO11 subgroups in the discovery cohort. Contrary to the previous embedding, each resulting 2D marker reflects a sample (n=491) and the same patient can be represented multiple times. To emphasize temporal structure, DSO was used as an input for PHATE, in addition to the 10 cytokine and tissue damage log concentrations. We again centered the data (mean 0) and apply standard scaling (variance 1). We then upweighted the time variable by a factor of √8 in distance computations in PHATE to better visualize time, as suggested in (*22*). The resulting PHATE embedding is colored by the average of the 10 standard scaled log concentrations used as input.

To visualize the evolution of the DSO11 clusters, we computed one multivariate linear regression on the samples of each cluster using DSO as the explanatory variable and the 10 log concentrations as a multivariate response. The linear model of each cluster was used to obtain continuous log concentration predictions for the DSO0-28 range. The resulting log concentration curves were projected onto the 2D precomputed longitudinal embedding using interpolation with existing samples, as implemented in the PHATE Python package. Two-stage bootstrap (see Statistical analyses) was used to visualize the confidence ellipses representing 3 standard deviations around the average.

### Statistical analyses

#### Statistical comparisons of single variables

The type of statistical test is specified in the figure legends. Given the size of the cohorts, we opted for conservative non-parametric tests. Mann-Whitney U test (MW) were performed on unpaired contrasts of interest (ex: within cluster 1, survivor vs deceased). If multiple MW were performed in a same panel, we first performed a Kruskal-Wallis (KW) test, then the MW was corrected for multiple comparisons with Dunn’s multiple comparison test. For the comparison of categorical values (demographics table, Table 1), we applied Chi^2^ test. For comparisons between three paired values (measurement of cytokine+ S-specific T cell response), we performed a Friedman test, with correction using Dunn’s multiple comparison test. Subjects with missing data (for example, who did not enough cells to perform stimulation with all three peptide pools) were excluded from both the panels and statistical analysis.

In the setting of pie charts, permutation tests (10 000 permutations) were calculated using the SPICE software (https://niaid.github.io/spice/). All other statistical tests were performed with Prism v9.5.0 (GraphPad). Statistical tests were considered two-sided and p<0.05 was considered significant (bolded in the panels).

#### Two-stage bootstrap

The two-stage bootstrap [sometimes called Hierarchical Bootstrap (*52*)] is a resampling method that accounts for intra-patient correlation in instances of repeated measures. For a sample containing *n* patients, possibly having repeated measures, it first creates a *bootstrap sample*, by randomly sampling with replacement *n* patients. This entails that patients may be sampled more than once, or even be missing from the bootstrap sample. Then, for a given patient in the bootstrap sample, the second stage consists in randomly sampling its observations with replacement, generating a patient sub-sample with the same number of observations the patient had in the original sample. Again, in this stage, some of the patient’s observations can be sampled multiple times, or missing, owing to the fact that sampling is done with replacement. All such patient sub-samples are aggregated into a full bootstrap sample. Typically, one generates a large amount of bootstrap samples and uses the distribution of the test statistic across these bootstrap samples as an estimate of the true underlying distribution, which can be hard, or even impossible, to derive analytically (*53*). This method was used to compare, among the four patient clusters of the discovery cohort, i) antibody kinetics; ii) AUC for vRNA, and iii) PHATE cytokine trajectories.

#### Model of RBD-specific IgG, IgM, and IgA kinetics

Kinetics of IgG, IgA and IgM antibody production were modeled using a logistic curve fit of log(antibody) ~ DSO with the *drm* function of the *drc* R package (*54*) set to the L.4 function (i.e. the 4-parameter logistic curve). While the lower limit of detection was that of the assay, we did not set any upper limit of detection; yielding in effect 3 parameters of estimation (the location parameter, the upper limit of detection, and the slope of the curve at the location parameter value). The pairwise differences between location parameters of these curves, i.e., the DSO at which 50% of maximal antibody production was reached, were used to compare clusters’ antibody production kinetics. 1000 two-stage bootstrap samples were used to obtain confidence intervals around the estimates of pairwise differences between location parameters of the logistic curves. The main strength of this testing approach is that we need not rely on strong assumptions to find a suitable distribution for the test statistic (i.e. the pairwise difference between the DSO at which 50% of maximal antibody production was reached), thanks to bootstrapping. As such, this method is robust to misspecification of the kinetics model.

#### Area Under Curve (AUC) on plasma viral RNA quantities over time

To quantify viral load among viremic patients, we estimate average Viral Load (copies/ml) * Time (DSO) as an Area Under Curve (AUC) for each cluster, the curves being Generalized Additive Models (GAM) with smooth spline estimations of the relationship Viral Load ~ Time over the period ranging from DSO0 to DSO30. We used the Lower Level of Quantification (LLOQ) threshold of 65 copies/ml. We chose to not transform Viral Load to allow for interpretation of the AUC based on the original units (copies/mL * time). We used the R package mgcv function gam. Four (4) knots and gaussian kernel smoothing gave the best bias-variance tradeoff. AUC was computed for DSO0 to DSO25 to eliminate inherent instability at border (DSO30) of smooth spline fits. The fitted values at DSO0 were all equal to the LLOQ threshold, so no such instability was present. AUC inter-cluster pairwise differences were computed for 1000 two-stage bootstrap simulations.

To only consider patients with RNAemia, patient observations retained were those with at least 1 measurement over the LLOQ threshold within the DSO0-30 timeframe. The results were robust to the presence of outlier patient 268 from CHUM.

**Table 3.**
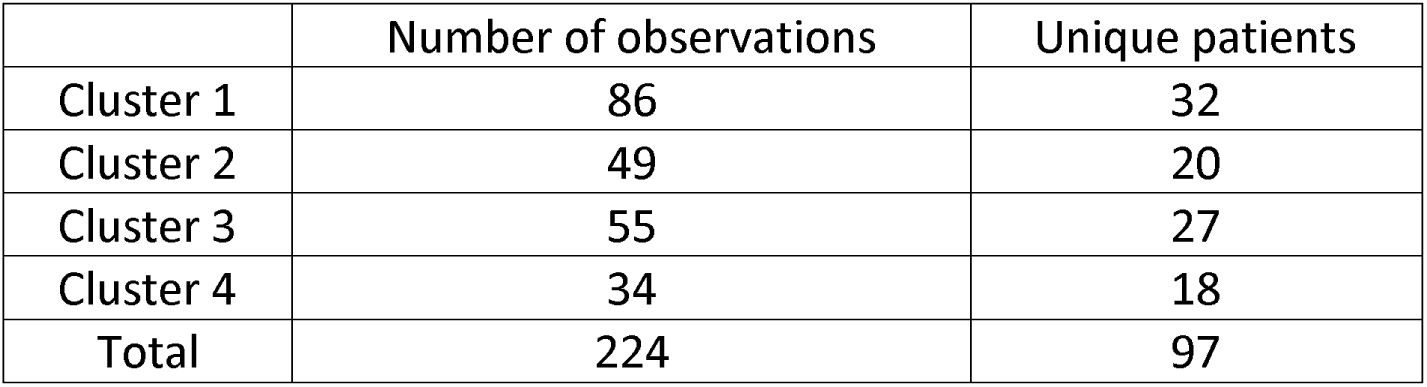
Number of viremic patients, with their respective sampling counts, in the discovery cohort.

|  | Number of observations | Unique patients |
| --- | --- | --- |
| Cluster 1 | 86 | 32 |
| Cluster 2 | 49 | 20 |
| Cluster 3 | 55 | 27 |
| Cluster 4 | 34 | 18 |
| Total | 224 | 97 |

#### Power calculation for the number of samples to test for RBD-specific B cells

We computed power curve estimates to determine the sample sizes necessary to assess the hypothesis of inter-cluster variation (Cluster 2 vs Cluster 1 and Cluster 4 vs Cluster 3) of RBD-specific B-cell counts (RBD.B) with appropriate power. No useful previous estimates of effect sizes were found in existing literature. To circumvent this lack of information, we used RBD-specific IgG cell counts (RBD.IgG) as a proxy of RBD.B, based on previous evidence of their association(*29*). Relationship between RBD.B and RBD.IgG was determined based on a linear regression fit on a sample of n=14 patients for which both measures were available; variability in this relationship was simulated using 1000 vanilla bootstrap simulations of the above sample. These 1000 regressions all at once were used to predict the values of RBD.B from RBD.IgG for 1000 distinct bootstrap samples of n=216 patients during the acute phase of the infection (~ DSO11). In each of these simulated samples, an effect size measure (Cohen’s d) was computed for RBD.B. We thus obtain a distribution of effect sizes for both inter-cluster differences, from which we computed proper sample size according to a conservative estimate based on 97.5^th^ percentile of the effect size distributions and a standard power of 0.8. The sample sizes obtained were n=32 and n=9 per cluster for inter-cluster differences Cluster 2 vs Clus er 1 and Cluster 4 vs Cluster 3, respectively.

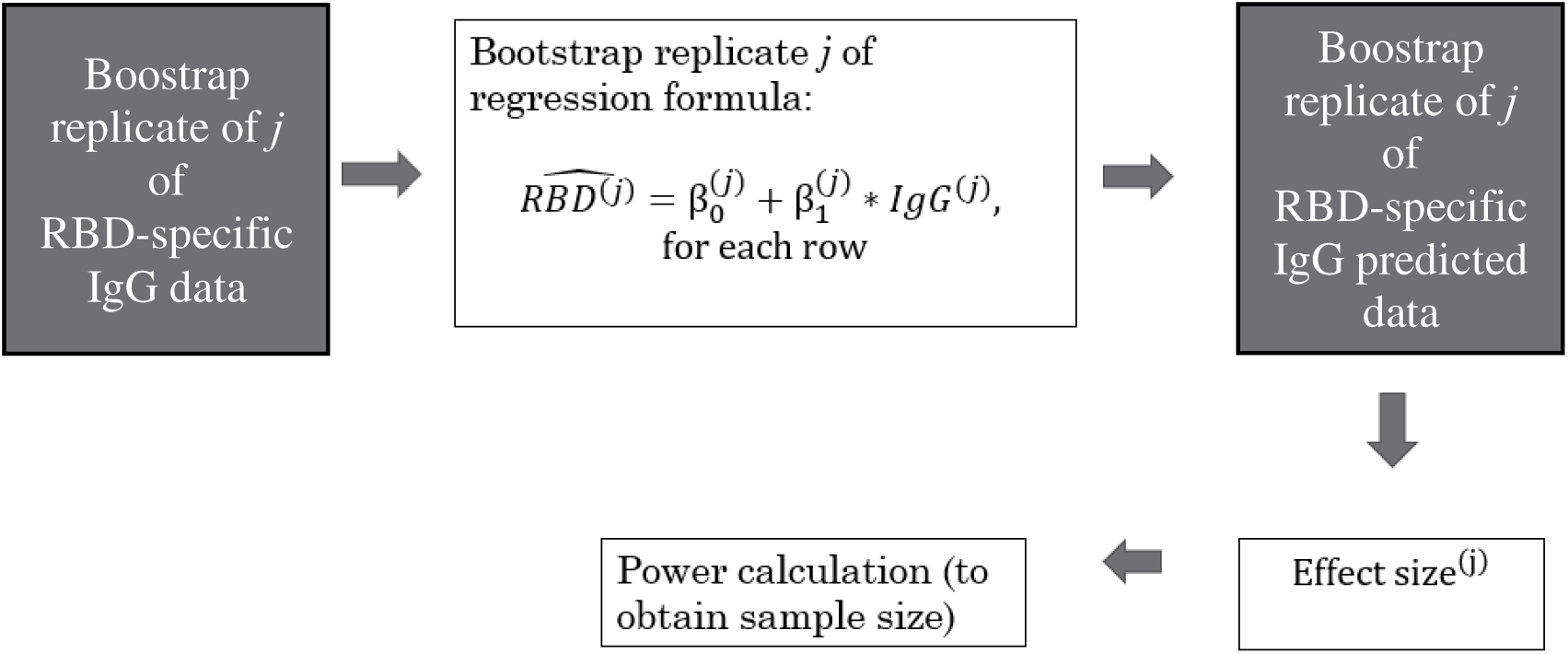

### Bulk RNA Sequencing

#### Sample Collection

Blood was collected into PAXgene Blood RNA tubes (BD Biosciences; San Jose, CA, USA) to ensure stabilization of intracellular RNA. Immediately after collection, tubes were inverted 10 times, kept at RT for 24hrs, −20°C for an additional 24hrs, and stored at −80°C. Batches of tubes were thawed overnight and total RNA was manually extracted using PAXgene Blood RNA Kit (Qiagen; Germantown, MD, USA), as per manufacturer instructions. Total RNA was quantified, and its integrity assessed on a LabChip GXII (PerkinElmer) instrument. Libraries were generated from 250 ng of total RNA as follows: mRNA enrichment was performed using the NEBNext Poly(A) Magnetic Isolation Module (New England BioLabs). cDNA synthesis was achieved with the NEBNext RNA First Strand Synthesis and NEBNext Ultra Directional RNA Second Strand Synthesis Modules (New England BioLabs). The remaining steps of library preparation were done using the NEBNext Ultra II DNA Library Prep Kit for Illumina (New England BioLabs). Adapters and PCR primers were purchased from New England BioLabs. Libraries were quantified using the Kapa Illumina GA with Revised Primers-SYBR Fast Universal kit (Kapa Biosystems). Average size fragment was determined using a LabChip GXII (PerkinElmer) instrument. The libraries were normalized and pooled and then denatured in 0.05N NaOH and neutralized using the HT1 buffer. The pool was loaded at 225pM on an Illumina NovaSeq S4 lane as per the manufacturer’s recommendations. The run was performed for 2×100 cycles (paired-end mode). A phiX library was used as a control and mixed with libraries at 1% level. Base calling was performed with RTA v3.4.4. Program bcl2fastq2 v2.20 was then used to demultiplex samples and generate fastq reads. The average base quality score for each sample dataset was verified to be Q33 or above and the % of aligned reads on reference sequence homo sapiens:hg19 was verified to be 90% or above.

#### Data processing and quality control

The sequencing reads were trimmed using CutAdapt (*55*) and mapped to the human reference genome (hg19) using 2STAR (*56*) aligner (version 2.6.1d), with default parameters. Only samples with bulk RNA-sequencing data and PHATE cluster assignments were kept for downstream analysis (n = 445). Expression data was filtered for protein-coding genes that were sufficiently expressed across all samples (median logCPM > 1, n = 10,236 genes retained after filtering). After removing non-coding and lowly-expressed genes, normalization factors to scale the raw library sizes were calculated using calcNormFactors in edgeR (v3.26.8) (*57*). The voom (*58*) function in limma (v3.40.6) was used to apply these size factors, estimate the mean-variance relationship, and convert counts to logCPM values. The technical effect of collection center (i.e., Centre Hospitalier de l’Université de Montréal vs Jewish General Hospital) was regressed using limma (v3.40.6) prior to downstream modeling.

#### Modeling PHATE effects

PHATE effects (i.e., the differential expression effects between individuals in PHATE clusters of interest) were modeled in individuals collected at the DSO11 timepoint with expression data (n = 174; n PHATE cluster 1 = 37, n PHATE cluster 2 = 35, n PHATE cluster 3 = 41, n PHATE cluster 4 = 61 individuals). All pairwise PHATE clusters contrasts were performed with the main contrasts of interest being cluster 1 vs 2, cluster 3 vs 4, and, within cluster 1 individuals, survivor (n = 19) vs deceased (n = 18). To obtain estimates of the PHATE effects, the following linear model was run for each pairwise PHATE contrast:

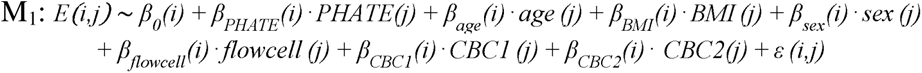

Here, *β_0_(i)* is the global intercept accounting for the expected collection center-corrected expression of gene *i* in a female individual in the baseline PHATE cluster, and *β_PHATE_(i)* indicates the effect of the non-baseline PHATE cluster (*PHATE(j)*) on gene *i*. For example, in the PHATE cluster contrast 1 vs 2, individuals in PHATE cluster 2 represent the baseline gene expression signature, and *β_PHATE_(i)* represents the effect of PHATE cluster 1 on gene expression. Further, age represents the mean-centered, scaled (mean = 0, sd = 1) age per individual, body mass index (BMI) represents the mean-centered, scaled (mean = 0, sd = 1) BMI per individual, sex represents the self-identified sex for each individual (factor levels = “Female”, “Male”), and flow cell represents the flow cell on which the sample was sequenced (seven factors in total). If BMI was not reported for an individual, this missing data was filled with the average BMI value across all individuals. Because we modeled whole blood expression data, two additional covariates were included, corresponding to the first two principal components of a PCA performed on an n x m cell type proportion matrix (where n = number of samples = 630, m = number of cell types = 5, with the matrix populated by the cell type proportions derived from clinical complete blood count [CBC] data) to account for the majority of the variance introduced by underlying cell type composition (PC1 percent variance explained (PVE) = 88.3%, PC2 PVE = 7.8%, total = 96.1%). Their corresponding effects on gene expression are represented by *β_CBC1_(i)* and *β_CBC2_(i)*. Finally, *ε (i,j)* represents the residuals for each gene *i*, individual *j* pair.

These models were fit using the lmFit and eBayes functions in limma(*59*), and the estimates of the PHATE effects *β_PHATE_(i)* were extracted across all genes along with their corresponding p-values. We controlled for false discovery rates (FDR) using an approach analogous to that of Storey and Tibshirani (*60, 61*), which makes no explicit assumptions regarding the distribution of the null model but instead derives it empirically. To obtain a null, we performed 100 permutations, where PHATE cluster label was permuted across individuals.

#### Calculation of ssGSEA scores

To construct the IFN and COVID-19 severity score metrics, we calculated single-sample Gene Set Enrichment Analysis (ssGSEA) scores using the Gene Set Variation Analysis (GSVA) package in R (v1.32.0) with default parameters and method = “ssgsea”(*62*). For the IFN ssGSEA score, the input genes were those belonging to the hallmark IFN gamma and alpha response pathways(*63*). For the COVID-19 severity ssGSEA score, the input genes were those previously described to be positively associated with increased COVID-19 susceptibility in peripheral blood mononuclear cells from COVID-19 patients(*23*).

#### Gene set enrichment analyses

Gene set enrichment analyses were performed using two independent methods, including fgsea (https://bioconductor.org/packages/release/bioc/html/fgsea.html) and ClueGO(*25*). The enrichment program specifications and the data in which they were used to assess enrichments are described below:

The R package fgsea (v1.10.1) was used to perform gene set enrichment analysis for the cluster 1 vs 2 effects, cluster 3 vs 4 effects, and cluster 1 survivor vs deceased effects using the H hallmark gene sets(*24*). T-statistics were obtained directly from the topTable function in limma(*59*). The background set of genes were those sufficiently expressed (i.e. passed the lowly-expressed gene filter threshold) in the whole blood expression data. The t-statistics were then ranked, and these pre-ranked t-statistics were used to perform the enrichment using fgsea with the following parameters: minSize = 15, maxSize = 500, nperm = 100000. Enrichment scores (ES) and Benjamini-Hochberg (*64*) adjusted p-values output by fgsea were collected for each analysis.

Additionally, we performed gene set enrichment analysis separately for genes upregulated in cluster 1 and cluster 3 individuals (i.e., the low antibody response clusters) relative to all other genes tested using ClueGO (v2.5.7) (*25*) in functional analysis mode. The target set of genes was the list of significantly upregulated genes in the cluster 1 or cluster 3 individuals (in the 1 vs 2 and 3 vs 4 contrasts, respectively) and the background set was the list of all genes tested. Specifically, we tested for the enrichment of GO terms related to biological processes (ontology source: GO_BiologicalProcess-EBI-UniProt-GOA_04.09.2018_00h00) using the following parameters: visual style = Groups, default Network Specificity, GO Term Fusion = TRUE, min. GO Tree Interval level = 3, max. GO Tree Interval level = 8, min. number of genes = 3, min. percentage of genes = 4.0, statistical test used = Enrichment (right-sided hypergeometric test), p-value correction = Bonferroni step down. For the graphical representation of the enrichment analysis, ClueGO clustering functionality was used (kappa threshold score for considering or rejecting term-to-term links set to 0.4). Only pathways with an FDR < 0.05 were reported.

Scripts and processed data: https://github.com/herandolph/COVID-19_PHATE

### Flow cytometry assessment of PBMCs

#### Antibodies and reagents

All antibodies are listed in Supplementary Tables 4 and 5. Antibodies are monoclonal and raised in mice or rats. All antibodies were validated by manufacturer and titrated with biological and/or isotype controls. SARS-CoV-2 Spike receptor binding domain (RBD) recombinant protein was expressed in Freestyle 293F cells and purified by nickel affinity columns, as directed by the manufacturer (Thermo Fisher Scientific). The RBD preparations were dialyzed against phosphate-buffered saline (PBS) and purity was assessed, by SDS-PAGE and Coomassie Blue staining. We generated B cell probes by conjugating recombinant RBD proteins with Alexa Fluor 488 dye or Alexa Fluor 594 dye (Thermo Fisher Scientific) according to the manufacturer’s protocol.

#### Detection of RBD-specific B cells

Cryopreserved peripheral blood mononuclear cells (PBMCs) were thawed and rested in cell culture media (RPMI supplemented with 10% fetal bovine serum (FBS) and PenStrep – 50 U/ml of penicillin and 50 µg/mL of streptomycin) at 37°C for 3hrs at a density of 1×10^7^ cells/ml in 24-well plates. Cells were collected, washed, and stained with LIVE/DEAD™ Fixable Aqua Dead Cell Stain Kit (20 mins, 4°C; Thermofisher, #L34965). After washing, cells were stained with a cocktail of surface markers (30 mins, 4°C; See panel in Supplementary Table 1, including RBD probes). Washed cells were then fixed with 2% paraformaldehyde (PFA) for 20 mins at RT, then washed and resuspended in PBS-2% FBS for flow acquisition on a 5-laser Symphony (BD). Analyses were performed using FlowJo (Treestar, V10).

#### Activation-induced marker (AIM) assay on T cells

Cryopreserved PBMCs were thawed and rested in cell culture media (RPMI supplemented with 10% Human AB serum and PenStrep – 50 U/mL of penicillin and 50 µg/mL of streptomycin) at 37°C for 3hrs at a density of 10M/mL in 24-well plates. 15 minutes prior to stimulation, CD40 blocking antibody (clone HB14, Miltenyi, cat #: 130-094-133) was added to each well at 0.5 µg/ml, as well as antibodies staining CXCR5, CXCR3 and CCR6. Cells were either left unstimulated or stimulated with overlapping peptide pools of Spike (S1 + S2), at a final concentration of 0.5 µg/mL/peptide. Alternatively, 1µg/ml of Staphylococcal Enterotoxin B (SEB, Toxin Technology) was used to stimulate the cells as a positive control. Cells were stimulated for 15hrs, collected, washed, and stained with LIVE/DEAD™ Fixable Aqua Dead Cell Stain Kit (20 mins, 4°C; Thermofisher, #L34965). After washing, cells were incubated with FcR block (10mins, 4°C; Miltenyi) then stained with a cocktail of surface markers (30 mins, 4°C; See panel in Supplementary Table 1). Washed cells were then fixed with 2% paraformaldehyde (PFA) for 20 mins at RT, then washed and resuspended in PBS-2% FBS for flow acquisition on a 5-laser Symphony (BD).

#### Intracellular cytokine staining (ICS) in Spike-specific T cells

Cryopreserved peripheral blood mononuclear cells (PBMCs) were thawed and rested for 2hrs in cell culture media. Cells were stimulated with overlapping peptide pools for SARS-CoV-2 spike (S), membrane (M) and nucleocapsid (NC) (0.5 μg/ml per peptide from JPT, Berlin, Germany) for 6hrs in the presence of anti-CD107a BV786 (BD Biosciences), Brefeldin A (BD Biosciences) and monensin-1 (BD Biosciences) at 37 °C and 5% CO_2_. DMSO-treated cells served as negative control and SEB-treated cells as positive control. Cells were stained with LIVE/DEAD™ Fixable Aqua Dead Cell Stain Kit (20 mins, 4°C; Thermofisher, #L34965) and surface markers (30 mins, 4°C), followed by detection of intracellular markers using the IC Fixation/Permeabilization kit (Thermo Fisher) according to the manufacturer’s protocol before acquisition at 5-laser Symphony (BD) (see Supplementary Table 6 for panel).

## Supporting information

Study participants - meta data

PHATE effects

DE enrichments

Flow cytometry panels

## Data Availability

Data from the Pronmed study is available from the SciLifeLab data repository after appropriate permissions and data access agreements (https://doi.org/10.17044/scilifelab.14229410). Data from the BQC-19 repository is available after appropriate permissions and data access agreements (https://en.quebeccovidbiobank.ca).

## Acknowledgment

The authors are grateful to the study participants and the clinical research teams. We thank the CRCHUM BSL3 and Flow Cytometry Platforms for technical assistance.

The Pronmed study acknowledges the clinical research team, particularly the research nurses Joanna Wessbergh and Elin Söderman and the biobank research assistants Erik Danielsson, Philip Karlsson, Labolina Spång and Amanda Svensson for their help in compiling the study.

## Authors contributions

<u>Conceptualization:</u> EBR, SM, HER, LB, GW, DEK

<u>Data Curation</u>: EBR, ML, AP, LM, MH, EB, SZ, TN, DM, JR, CL, AP, NA

<u>Formal analysis</u>: EBR, SM, HER, ML, JB, RLB, AP, LM, MH

<u>Funding acquisition</u>: DEK, GW, LB, MT, AF, NC, JBR, MT

<u>Investigation/experiments:</u> EBR, AP, LM, MH, JN, RC, AS, MB, JP, SD, SPA, ML, RF, AL, DV, HM, FP

<u>Biostatistical methodology:</u> JB, RLB, HR

<u>Patient recruitment and cohort administration:</u> NB, MD, DM, NA, CL, AP, JBR, DEK

<u>Visualization:</u> EBR, SM, HER, JB, RLB, GS

<u>Supervision:</u> MT, NC, LB, KM, GW, JR, AF, DEK

<u>Writing – original draft:</u> EBR, SM, HER, DEK

<u>Writing – review and editing:</u> all authors

## Competing interests

J.B.R. has served as an advisor to GlaxoSmithKline and Deerfield Capital. TN has received speaking fee from Boehringer Ingelheim for the talks unrelated to this research. These agencies had no role in the design, implementation, or interpretation of this study. The authors declare that they have no other competing interests.

## Funding

This study was funded by the American Foundation for AIDS Research (amfAR) grant 110068-68-RGCV (DEK, NC, AF); Canada’s COVID-19 Immunity Task Force (CITF), in collaboration with the Canadian Institutes of Health Research (CIHR) grant VR2-173203 (DEK, AF); CIHR grant # 178344 (DEK, AF); CIHR grants 365825 and 409511 (JBR); Canada Foundation for Innovation (CFI): Exceptional Fund COVID-19 grant #41027 to AF, DEK, NC; CFI leader to JBR. The Symphony flow cytometer was funded by a John R. Evans Leaders Fund from the Canada Foundation for Innovation (#37521 to D.E.K) and the Fondation Sclérodermie Québec. The Ministère de l’Économie et de l’Innovation du Québec, Programme de soutien aux organismes de recherche et d’innovation (AF), CRCHUM Foundation. The Biobanque Québécoise de la COVID-19 (BQC-19) is supported by the Fonds de recherche Québec-Santé (FRQS) Génome Québec and the Public Health Agency of Canada. DEK and JBR are FRQS Merit Research Scholars. NC, MD, MC, JBR, CL and MT are supported by FRQS Salary Awards. AF and AP are recipients of Canada Research Chairs. EBR is recipient of a COVID-19 excellence scholarship from the Université de Montréal (EBR); SM is recipient of an IVADO MSc Excellence scholarship and a Fonds de recherche du Québec – Nature et technologies (FRQNT) B1X scholarship (SM); HER is supported by a National Institutes of Health (NIH) Ruth L. Kirschstein National Research Service Award (F31-HL156419); SPA, J.P., M.B. were supported by CIHR fellowships; G.S is supported by a FRQS doctoral fellowship and by a scholarship from the Department of Microbiology, Infectious Disease, and Immunology of the University of Montreal.

The Pronmed study was funded by the SciLifeLab/Knut and Alice Wallenberg national COVID-19 research program (M.H.: KAW 2020.0182, KAW 2020.0241), the Swedish Heart-Lung Foundation (M.H.: 20210089, 20190639, 20190637), the Swedish Research Council (R.F.: 2014-02569, 2014-07606), the Swedish Kidney Foundation (R.F.: F2020-0054) and the Swedish Society of Medicine (M.H.: SLS-938101). Funding bodies had no role in the design of the study, data collection, interpretation, or in the writing of the manuscript.

## Materials & Correspondence

Please direct correspondence relating to this article, as well as inquiries about data regarding the discovery cohort, to. For data relating to the validation cohort, please direct inquiries to.

## Notes

### Author Declarations

For participants enrolled in Quebec, the study was approved by the IRBs of the University of Montreal Hospital and the Jewish General Hospital (multicentric protocol: MP-02-2020-8929) and written informed consent obtained from all participants or, when incapacitated, their legal guardian before enrollment and sample collection. Research adhered to the standards indicated by the Declaration of Helsinki. For the validation cohort recruited at Uppsala University Hospital in Sweden, the study was approved by the Swedish National Ethical Review Agency (Pronmed study; 2017-043, amended 2019-00169, 2020-01623, 2020-05730 and 2022-00526-01) and registered a priori at ClinicalTrials.gov (NCT03720860). Informed consent was obtained from the patient or next of kin if the patient was unable to give consent. The Declaration of Helsinki and subsequent revisions were followed.

## REFERENCES

1. E. Brunet-Ratnasingham et al., Integrated immunovirological profiling validates plasma SARS-CoV-2 RNA as an early predictor of COVID-19 mortality. Sci Adv 7, eabj5629 (2021).

2. E. Pujadas et al., SARS-CoV-2 viral load predicts COVID-19 mortality. Lancet Respir Med 8, e70 (2020).

3. J. D. Jarhult, M. Hultstrom, A. Bergqvist, R. Frithiof, M. Lipcsey, The impact of viremia on organ failure, biomarkers and mortality in a Swedish cohort of critically ill COVID-19 patients. Sci Rep 11, 7163 (2021).

4. R. C. Group, Baricitinib in patients admitted to hospital with COVID-19 (RECOVERY): a randomised, controlled, open-label, platform trial and updated meta-analysis. Lancet 400, 359–368 (2022).

5. R. Rovito et al., Association between SARS-CoV-2 RNAemia and dysregulated immune response in acutely ill hospitalized COVID-19 patients. Sci Rep 12, 19658 (2022).

6. S. Bulow Anderberg et al., Increased levels of plasma cytokines and correlations to organ failure and 30-day mortality in critically ill Covid-19 patients. Cytokine 138, 155389 (2021).

7. A. G. Laing et al., A dynamic COVID-19 immune signature includes associations with poor prognosis. Nat Med 26, 1623–1635 (2020).

8. C. Lucas et al., Longitudinal analyses reveal immunological misfiring in severe COVID-19. Nature 584, 463–469 (2020).

9. R. C. Group, Tocilizumab in patients admitted to hospital with COVID-19 (RECOVERY): a randomised, controlled, open-label, platform trial. Lancet 397, 1637–1645 (2021).

10. R.-C. Investigators et al., Interleukin-6 Receptor Antagonists in Critically Ill Patients with Covid-19. N Engl J Med 384, 1491–1502 (2021).

11. R. C. Group et al., Dexamethasone in Hospitalized Patients with Covid-19. N Engl J Med 384, 693–704 (2021).

12. P. Bastard et al., Autoantibodies against type I IFNs in patients with life-threatening COVID-19. Science 370, (2020).

13. Q. Zhang et al., Inborn errors of type I IFN immunity in patients with life-threatening COVID-19. Science 370, (2020).

14. W. H. O. S. T. Consortium et al., Repurposed Antiviral Drugs for Covid-19 - Interim WHO Solidarity Trial Results. N Engl J Med 384, 497–511 (2021).

15. A. C. Kalil et al., Efficacy of interferon beta-1a plus remdesivir compared with remdesivir alone in hospitalised adults with COVID-19: a double-bind, randomised, placebo-controlled, phase 3 trial. Lancet Respir Med 9, 1365–1376 (2021).

16. R. Nienhold et al., Two distinct immunopathological profiles in autopsy lungs of COVID-19. Nat Commun 11, 5086 (2020).

17. S. Asif et al., Weak anti-SARS-CoV-2 antibody response is associated with mortality in a Swedish cohort of COVID-19 patients in critical care. Crit Care 24, 639 (2020).

18. T. Zohar et al., Compromised Humoral Functional Evolution Tracks with SARS-CoV-2 Mortality. Cell 183, 1508–1519 e1512 (2020).

19. C. Rydyznski Moderbacher et al., Antigen-Specific Adaptive Immunity to SARS-CoV-2 in Acute COVID-19 and Associations with Age and Disease Severity. Cell 183, 996–1012 e1019 (2020).

20. P. S. Arunachalam et al., Systems biological assessment of immunity to mild versus severe COVID-19 infection in humans. Science 369, 1210–1220 (2020).

21. C. Liu et al., Time-resolved systems immunology reveals a late juncture linked to fatal COVID-19. Cell 184, 1836–1857.e1822 (2021).

22. K. R. Moon et al., Visualizing structure and transitions in high-dimensional biological data. Nat Biotechnol 37, 1482–1492 (2019).

23. H. E. Randolph et al., Genetic ancestry effects on the response to viral infection are pervasive but cell type specific. Science 374, 1127–1133 (2021).

24. A. Subramanian et al., Gene set enrichment analysis: a knowledge-based approach for interpreting genome-wide expression profiles. Proc Natl Acad Sci U S A 102, 15545–15550 (2005).

25. G. Bindea et al., ClueGO: a Cytoscape plug-in to decipher functionally grouped gene ontology and pathway annotation networks. Bioinformatics 25, 1091–1093 (2009).

26. D. Mathew et al., Deep immune profiling of COVID-19 patients reveals distinct immunotypes with therapeutic implications. Science 369, (2020).

27. D. G. Brooks, L. Teyton, M. B. Oldstone, D. B. McGavern, Intrinsic functional dysregulation of CD4 T cells occurs rapidly following persistent viral infection. J Virol 79, 10514–10527 (2005).

28. J. R. Teijaro et al., Persistent LCMV infection is controlled by blockade of type I interferon signaling. Science 340, 207–211 (2013).

29. M. Nayrac et al., Temporal associations of B and T cell immunity with robust vaccine responsiveness in a 16-week interval BNT162b2 regimen. Cell Rep 39, 111013 (2022).

30. J. Fajnzylber et al., SARS-CoV-2 viral load is associated with increased disease severity and mortality. Nat Commun 11, 5493 (2020).

31. R. L. Gottlieb et al., Early Remdesivir to Prevent Progression to Severe Covid-19 in Outpatients. N Engl J Med 386, 305–315 (2022).

32. D. M. Weinreich et al., REGEN-COV Antibody Combination and Outcomes in Outpatients with Covid-19. N Engl J Med 385, e81 (2021).

33. A.-T. L.-C. S. Group et al., A Neutralizing Monoclonal Antibody for Hospitalized Patients with Covid-19. N Engl J Med 384, 905–914 (2021).

34. R. C. Group, Casirivimab and imdevimab in patients admitted to hospital with COVID-19 (RECOVERY): a randomised, controlled, open-label, platform trial. Lancet 399, 665–676 (2022).

35. S. Crotty, Follicular helper CD4 T cells (TFH). Annu Rev Immunol 29, 621–663 (2011).

36. J. Major et al., Type I and III interferons disrupt lung epithelial repair during recovery from viral infection. Science 369, 712–717 (2020).

37. J. D. Domizio et al., The cGAS-STING pathway drives type I IFN immunopathology in COVID-19. Nature 603, 145–151 (2022).

38. E. B. Wilson et al., Blockade of chronic type I interferon signaling to control persistent LCMV infection. Science 340, 202–207 (2013).

39. T. N. Hoang et al., Modulation of type I interferon responses potently inhibits SARS-CoV-2 replication and inflammation in rhesus macaques. bioRxiv, (2022).

40. M. Papatriantafyllou, The interferon paradox. Nature Reviews Immunology 13, 392–393 (2013).

41. N. G. Sandler et al., Type I interferon responses in rhesus macaques prevent SIV infection and slow disease progression. Nature 511, 601–605 (2014).

42. E. Pairo-Castineira et al., Genetic mechanisms of critical illness in COVID-19. Nature 591, 92–98 (2021).

43. J. S. Lee et al., Immunophenotyping of COVID-19 and influenza highlights the role of type I interferons in development of severe COVID-19. Sci Immunol 5, (2020).

44. D. Blanco-Melo et al., Imbalanced Host Response to SARS-CoV-2 Drives Development of COVID-19. Cell 181, 1036–1045 e1039 (2020).

45. D. Jhuti et al., Interferon Treatments for SARS-CoV-2: Challenges and Opportunities. Infect Dis Ther 11, 953–972 (2022).

46. J. Prevost et al., Cross-Sectional Evaluation of Humoral Responses against SARS-CoV-2 Spike. Cell Rep Med 1, 100126 (2020).

47. M. Yuan et al., A highly conserved cryptic epitope in the receptor binding domains of SARS-CoV-2 and SARS-CoV. Science 368, 630–633 (2020).

48. K. Pearson, LIII. On lines and planes of closest fit to systems of points in space. *The London*, Edinburgh, and Dublin Philosophical Magazine and Journal of Science 2, 559–572 (1901).

49. S. P. Lloyd, Least squares quantization in PCM. IEEE Trans. Inf. Theory 28, 129–136 (1982).

50. F. Pedregosa et al., Scikit-learn: Machine learning in Python. the Journal of machine Learning research 12, 2825–2830 (2011).

51. D. B. Burkhardt et al., Enhancing experimental signals in single-cell RNA-sequencing data using graph signal processing. (2019).

52. V. Saravanan, G. J. Berman, S. J. Sober, Application of the hierarchical bootstrap to multi-level data in neuroscience. Neuron Behav Data Anal Theory 3, (2020).

53. B. Efron, R. J. Tibshirani, An Introduction to the Bootstrap. (CRC Press, 1994).

54. C. Ritz, F. Baty, J. C. Streibig, D. Gerhard, Dose-Response Analysis Using R. PLoS One 10, e0146021 (2015).

55. M. Martin, Cutadapt removes adapter sequences from high-throughput sequencing reads. EMBNET.journal 17, 10–12 (2011).

56. A. Dobin et al., STAR: ultrafast universal RNA-seq aligner. Bioinformatics 29, 15–21 (2013).

57. M. D. Robinson, D. J. McCarthy, G. K. Smyth, edgeR: a Bioconductor package for differential expression analysis of digital gene expression data. Bioinformatics 26, 139–140 (2010).

58. C. W. Law, Y. Chen, W. Shi, G. K. Smyth, voom: precision weights unlock linear model analysis tools for RNA-seq read counts. Genome Biology 15, R29 (2014).

59. M. E. Ritchie et al., limma powers differential expression analyses for RNA-sequencing and microarray studies. Nucleic Acids Res 43, e47 (2015).

60. Y. Nedelec et al., Genetic Ancestry and Natural Selection Drive Population Differences in Immune Responses to Pathogens. Cell 167, 657–669 e621 (2016).

61. J. D. Storey, R. Tibshirani, Statistical significance for genomewide studies. Proc Natl Acad Sci U S A 100, 9440–9445 (2003).

62. S. Hanzelmann, R. Castelo, J. Guinney, GSVA: gene set variation analysis for microarray and RNA-seq data. BMC Bioinformatics 14, 7 (2013).

63. A. Liberzon et al., The Molecular Signatures Database (MSigDB) hallmark gene set collection. Cell Syst 1, 417–425 (2015).

64. Y. Hochberg, Y. Benjamini, More powerful procedures for multiple significance testing. Stat Med 9, 811–818 (1990).

