## Supplemental legends and figures for "Sustained IFN signaling is associated with delayed development of SARS-CoV-2-specific immunity"

**SUPPLEMENTARY FIGURE LEGENDS**

**Supplementary Figure 1. Demographics are not differentially associated with patient clusters. AB**) At DSO11, plasma concentration across four patient clusters of RBD-specific **A**) IgM or **B**) IgA. **CD**) MELD representations of local enrichment across PHATE embedding, of C) critical severity or D) fatal outcome. **E**) Days in hospital among survivors of patient clusters. **F)** Age of patients per patient cluster. **G**) Percentage of the whole cohort or per patient cluster which are female (hashed). **H**) Fraction of dominant ancestry per patient cluster, calculated from genotyping. N discovery cohort: 1 = 38; 2 = 49; 3 = 73; 4 = 82 (242 in total). ABEF) Kruskal-Wallis with Dunn's multiple comparison tests. GH) Chi2 tests.

**Supplementary Figure 2. Kinetics of RBD-specific antibody responses per patient clusters. A)** RBD-specific IgG levels at DSO20 (+/- 4 days) per patient cluster. **B)** Sigmoidal curve fitted to the average per day per patient cluster of RBD-specific IgG responses, along day since symptom onset, within the validation cohort. **C)** RBD-specific IgG levels at DSO20 (+/- 4 days) per patient cluster of the validation cohort. **DE)** Model and statistical comparisons of RBD-specific D) IgM and E) IgA of the discovery cohort. **F**) Fraction of patients per cluster with at least one sample with detectable plasma vRNA (>13 copies/mL, saturated pie slices) throughout hospital stay. N discovery cohort: 1 = 38; 2 = 49; 3 = 73; 4 = 82 (242 in total). N validation cohort: V1 = 37; V2 = 39 (76 in total). A) Kruskal-Wallis with Dunn's multiple comparison tests. B) Mann-Whitney test. DE) Two-stage bootstrap, with 1000 simulations. Pairwise comparisons between all four clusters.

**Supplementary Figure 3. Similar plasma profiles at DSO11 between different outcomes of cluster 1 patients. A)** Patients from PHATE embedding color-coded by outcome at DSO60, where mauve dots are fatal outcome, and pink dots are survivors. **B-F)** DSO11 levels, between survivors and non-survivors, of **B**) plasma vRNA; **C**) Cytokine score ; **D**) Tissue damage score and **E**) RBD-specific IgG ; F) ssGSEA COVID severity score. **G**) GSEA on Hallmark gene sets for the contrast survivor vs deceased within cluster 1 only. **H-I)** Gene ontology (GO) enrichments for the Biological Processes (BP) database for significantly upregulated (FDR < 0.01, logFC > 0) genes for contrasts H) 1 vs 2 or I) 3 vs 4. Only significant enrichments (FDR < 0.05) are shown. **J-L**) At DSO11, correlation between IFN score and contemporaneous **J**) vRNA; **K**) Cytokine Score or **L**) Tissue damage score. BCDEFJKL) N: 1 = 38; 2 = 49 ; 3 = 73, 4 = 82 (242 in total). GHI). N: 1 = 37 ; 2 = 35 ; 3 = 41, 4 = 61 (174 in total).BCDEF) Mann-Whitney test. JKL) Spearman correlation.

**Supplementary Figure 4. Distribution of isotypes among RBD-specific antibody secreting cells. A)** Representative gating strategy to identify B cells (CD19+ CD20+) or plasmablasts (CD19+/- CD20- CD11c- CD38+ CD27+). **B)** Representative gating strategy of RBD-specific PB per cluster. **C)** RBD gate on B cells from a convalescent subject. **D)** Frequency of B cells which are RBD-specific in all four acute COVID-19 clusters, uninfected donors (UC) or convalescent subjects (Conv). **E)** RBD gate on plasma cells from a convalescent subject. **F)** Frequency of PB cells which are RBD-specific in all four acute COVID-19 clusters, uninfected donors (UC), or convalescent subjects (Conv). n for cluster 1 = 14; 2 = 16; 3 = 12; 4 = 13; Conv = 3; UC = 5. **GH)** Correlation between ssGSEA COVID-19 severity score and absolute counts of RBD-specific G) B cells or H) PB. **IJ)** Pie chart of isotype expression among I) RBD-specific B cells or J) RBD-specific PB per patient clusters. n for cluster 1 = 14; 2 = 16; 3 = 12; 4 = 13. DF) Kruskal-Wallis with Dunn's multiple comparison tests. GH) Spearman correlations. IJ) Permutation test (1000 iterations).

**Supplementary Figure 5. Spike-specific T cell responses in acute and resolved COVID-19. A)** PBMCs from acutely-SARS-CoV-2-infected individuals were stimulated with peptide pools of viral antigens (Spike – S , M or N) for 6hrs (addition of BFA 1 hour after stimulation). Representative gates on cytokine+ $CD4^+$ or $CD8^+$ T cells. **B)** Net frequency of SARS-CoV-2-specific $CD4^+$ T cells, as calculated using a Boolean OR gate on CD69 vs CD40L, IFNγ, IL-2, IL-10, IL-17, or TNFα. **C)** Net frequency of SARS-CoV-2-specific $CD8^+$ T cells, as calculated using a Boolean OR gate on CD69 vs

CD107a, IFNγ or TNFα. **D**) Following 9hrs stimulation of PBMCs with Spike peptide pool, representative gating strategy for identify $CD4^+$ or $CD8^+$ T cells using AIM panel. **EF)** Frequency of **E**) $CD4^+$ T or **F**) $CD8^+$ T cells which are Spike-specific in all four acute COVID-19 clusters, uninfected donors (UC), or convalescent subjects (Conv). C) n = 26 COVID-19+ acute samples (3 were excluded due to incomplete sampling). EF) n for cluster 1 = 11; 2 = 13; 3 = 9; 4 = 11; Conv = 5; UC = 5. C) Friedman test, with Dunn's multiple comparison test. EF) Kruskal-Wallis with Dunn's multiple comparison tests.

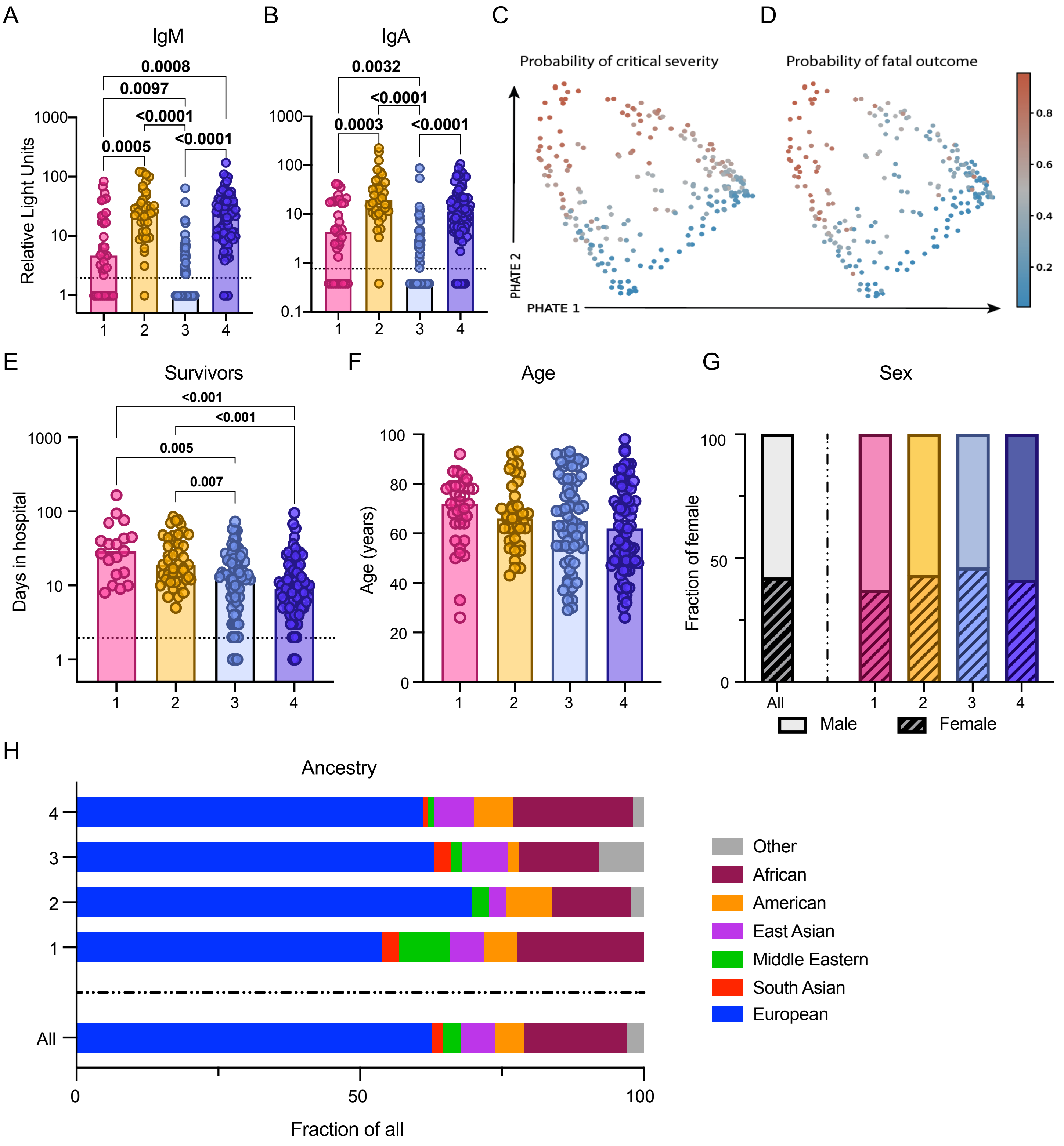
A
IgM
0.0008
0.0097
<0.0001
0.0005
<0.0001
Relative Light Units
1000
100
10
1
1
2
3
4
B
IgA
0.0032
<0.0001
0.0003
<0.0001
1000
100
10
1
0.1
C
Probability of critical severity
D
Probability of fatal outcome
PHATE 2
PHATE 1
0.8
0.6
0.4
0.2
E
Survivors
<0.001
<0.001
0.005
0.007
Days in hospital
F
Age
Age (years)
G
Sex
Fraction of female
All
Male
Female
H
Ancestry
Other
African
American
East Asian
Middle Eastern
South Asian
European
Fraction of all

S1

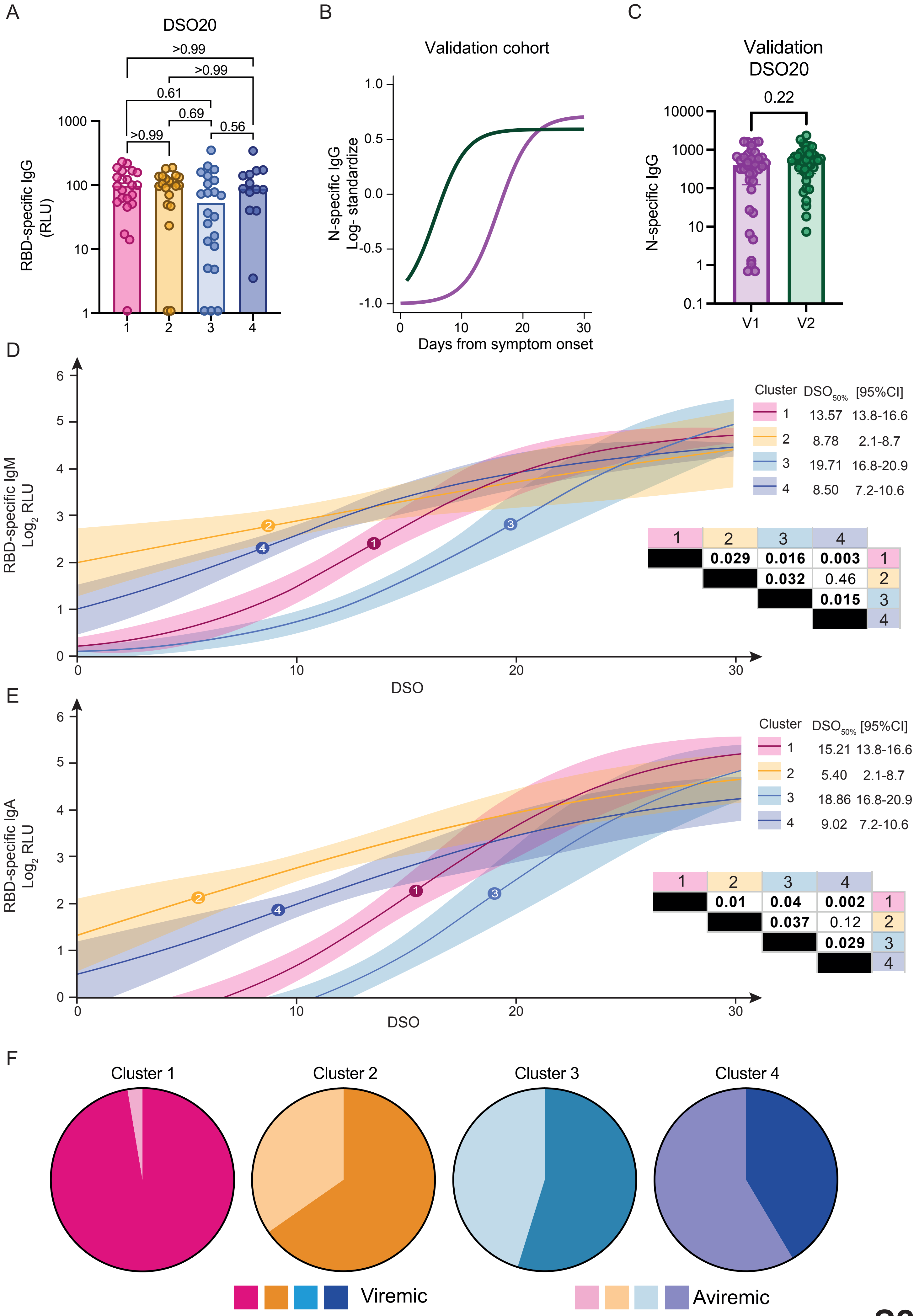
A
DSO20
>0.99
>0.99
0.61
0.69
>0.99
0.56
RBD-specific IgG (RLU)
1000
100
10
1
1
2
3
4
B
Validation cohort
N-specific IgG Log- standardize
1.0
0.5
0.0
-0.5
-1.0
0
10
20
30
Days from symptom onset
C
Validation DSO20
0.22
N-specific IgG
10000
1000
100
10
1
0.1
V1
V2
D
RBD-specific IgM Log2 RLU
Cluster DSO50% [95%CI]
1 13.57 13.8-16.6
2 8.78 2.1-8.7
3 19.71 16.8-20.9
4 8.50 7.2-10.6
1 2 3 4
0.029 0.016 0.003 1
0.032 0.46 2
0.015 3
4
DSO
E
RBD-specific IgA Log2 RLU
Cluster DSO50% [95%CI]
1 15.21 13.8-16.6
2 5.40 2.1-8.7
3 18.86 16.8-20.9
4 9.02 7.2-10.6
1 2 3 4
0.01 0.04 0.002 1
0.037 0.12 2
0.029 3
4
DSO
F
Cluster 1
Cluster 2
Cluster 3
Cluster 4
Viremic
Aviremic

**S2**

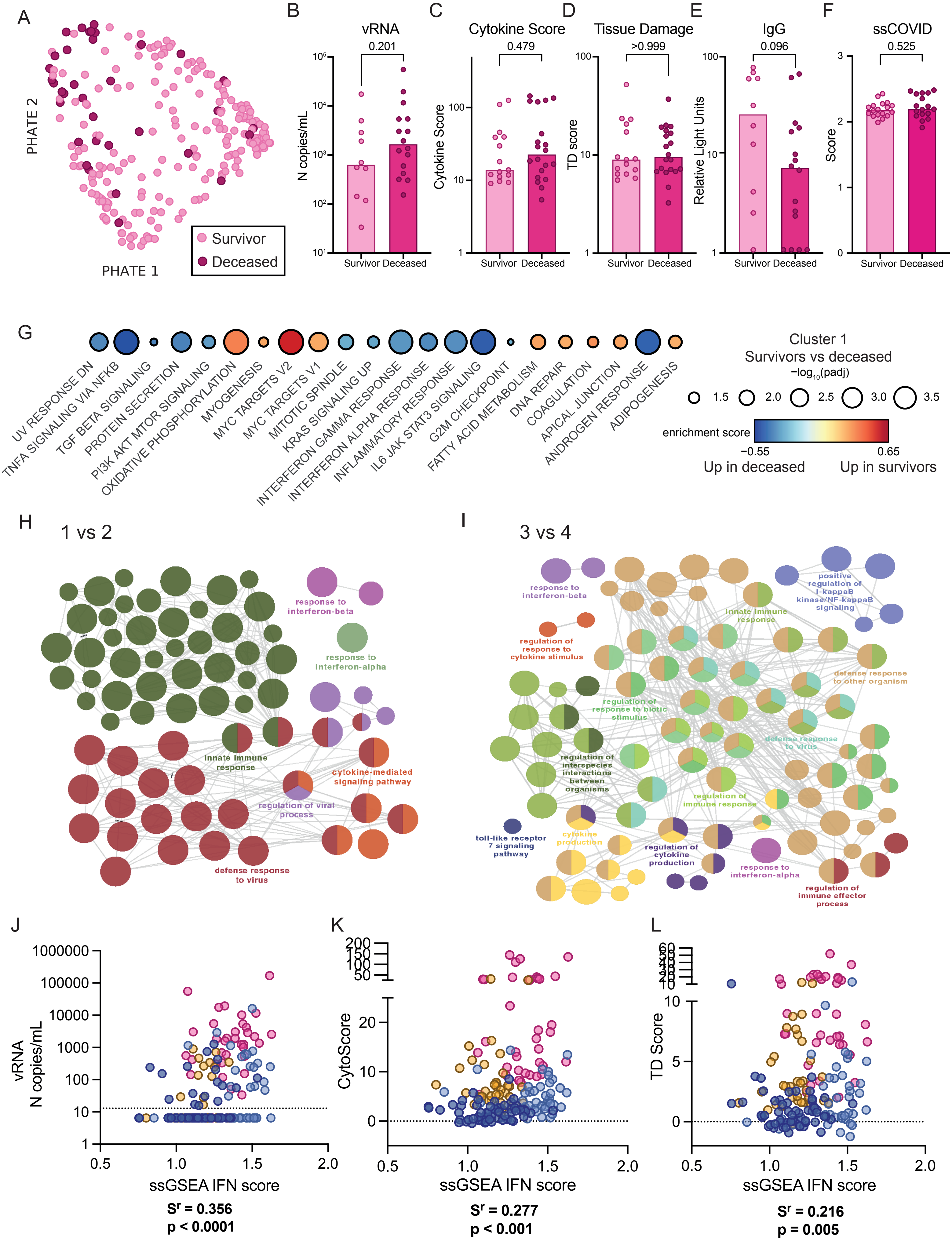
A
PHATE 2
PHATE 1
Survivor
Deceased
B
vRNA
0.201
N copies/mL
Survivor Deceased
C
Cytokine Score
0.479
Cytokine Score
Survivor Deceased
D
Tissue Damage
>0.999
TD score
Survivor Deceased
E
IgG
0.096
Relative Light Units
Survivor Deceased
F
ssCOVID
0.525
Score
Survivor Deceased
G
UV RESPONSE DN
TNFA SIGNALING VIA NFKB
TGF BETA SIGNALING
PROTEIN SECRETION
PI3K AKT MTOR SIGNALING
OXIDATIVE PHOSPHORYLATION
MYOGENESIS
MYC TARGETS V2
MYC TARGETS V1
MITOTIC SPINDLE
KRAS SIGNALING UP
INTERFERON GAMMA RESPONSE
INTERFERON ALPHA RESPONSE
INFLAMMATORY RESPONSE
IL6 JAK STAT3 SIGNALING
G2M CHECKPOINT
FATTY ACID METABOLISM
DNA REPAIR
COAGULATION
APICAL JUNCTION
ANDROGEN RESPONSE
ADIPOGENESIS
Cluster 1
Survivors vs deceased
−log10(padj)
1.5 2.0 2.5 3.0 3.5
enrichment score
−0.55 0.65
Up in deceased Up in survivors
H
1 vs 2
response to interferon-beta
response to interferon-alpha
innate immune response
cytokine-mediated signaling pathway
regulation of viral process
defense response to virus
I
3 vs 4
response to interferon-beta
positive regulation of I-kappaB kinase/NF-kappaB signaling
innate immune response
regulation of response to cytokine stimulus
defense response to other organism
regulation of response to biotic stimulus
defense response to virus
regulation of interspecies interactions between organisms
regulation of immune response
toll-like receptor 7 signaling pathway
cytokine production
regulation of cytokine production
response to interferon-alpha
regulation of immune effector process
J
vRNA N copies/mL
ssGSEA IFN score
Sr = 0.356
p < 0.0001
K
CytoScore
ssGSEA IFN score
Sr = 0.277
p < 0.001
L
TD Score
ssGSEA IFN score
Sr = 0.216
p = 0.005
S3

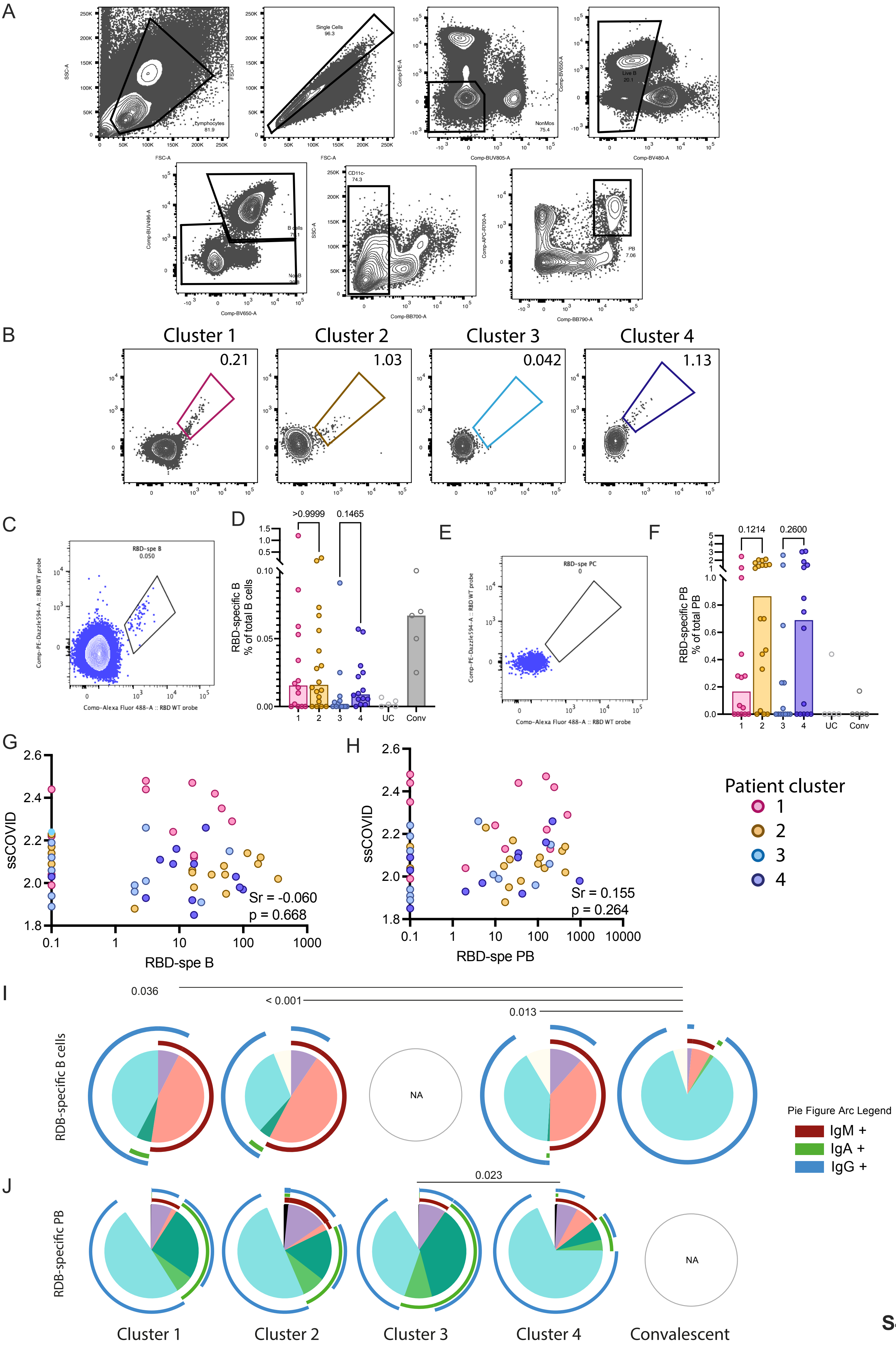
A
Lymphocytes 81.9
Single Cells 96.3
NonMos 75.4
Live B 20.1
B cells 73.1
NonB 20.8
CD11c- 74.3
PB 7.06
SSC-A
FSC-A
FSC-H
Comp-PE-A
Comp-BUV805-A
Comp-BV650-A
Comp-BV480-A
Comp-BUV496-A
Comp-BB700-A
Comp-APC-R700-A
Comp-BB790-A
B
Cluster 1
Cluster 2
Cluster 3
Cluster 4
0.21
1.03
0.042
1.13
C
RBD-spe B 0.050
Comp-PE-Dazzle594-A :: RBD WT probe
Comp-Alexa Fluor 488-A :: RBD WT probe
D
>0.9999
0.1465
RBD-specific B % of total B cells
1 2 3 4 UC Conv
E
RBD-spe PC 0
Comp-PE-Dazzle594-A :: RBD WT probe
Comp-Alexa Fluor 488-A :: RBD WT probe
F
0.1214
0.2600
RBD-specific PB % of total PB
1 2 3 4 UC Conv
G
ssCOVID
RBD-spe B
Sr = -0.060
p = 0.668
H
ssCOVID
RBD-spe PB
Sr = 0.155
p = 0.264
Patient cluster
1
2
3
4
I
0.036
< 0.001
0.013
RDB-specific B cells
NA
Pie Figure Arc Legend
IgM +
IgA +
IgG +
J
0.023
RDB-specific PB
NA
Cluster 1
Cluster 2
Cluster 3
Cluster 4
Convalescent

S4

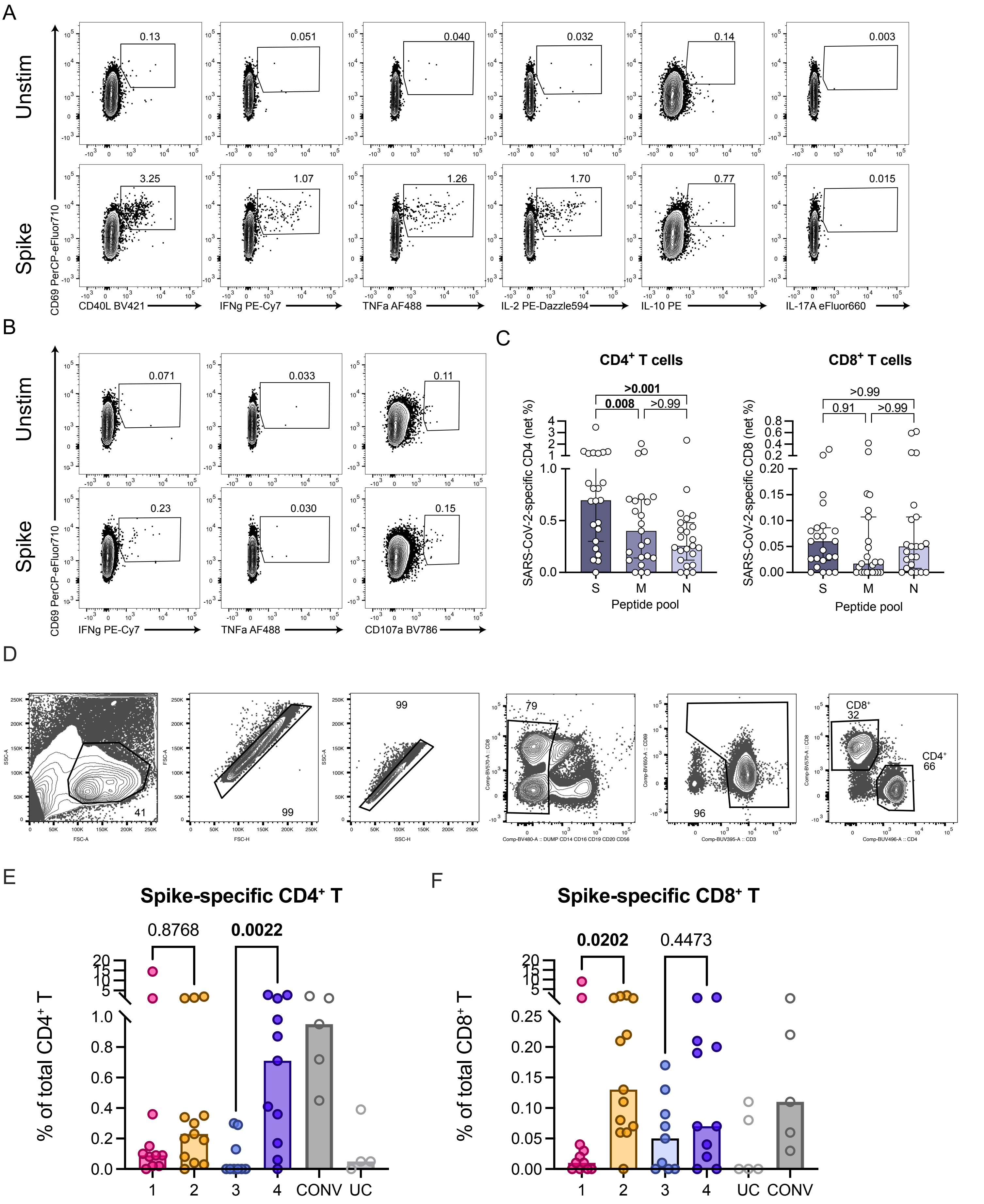
A
Unstim
Spike
CD69 PerCP-eFluor710
CD40L BV421
IFNg PE-Cy7
TNFa AF488
IL-2 PE-Dazzle594
IL-10 PE
IL-17A eFluor660
0.13
0.051
0.040
0.032
0.14
0.003
3.25
1.07
1.26
1.70
0.77
0.015
B
CD107a BV786
0.071
0.033
0.11
0.23
0.030
0.15
C
CD4+ T cells
CD8+ T cells
SARS-CoV-2-specific CD4 (net %)
SARS-CoV-2-specific CD8 (net %)
Peptide pool
>0.001
0.008
>0.99
>0.99
0.91
>0.99
S
M
N
D
E
Spike-specific CD4+ T
0.8768
0.0022
% of total CD4+ T
1
2
3
4
CONV
UC
F
Spike-specific CD8+ T
0.0202
0.4473
% of total CD8+ T

**S5**
