## Supplementary material for "Sustained IFN signaling is associated with delayed development of SARS-CoV-2-specific immunity": Flow cytometry panels

**Supplementary Table 4: Flow cytometry panel used to detect RBD-specific B and PC**

| Antigen/Reagent | Fluorochrome | Clone | Manufacturer | Cat # | Volume/test (µL)* |
| --- | --- | --- | --- | --- | --- |
| Brilliant Stain buffer | - | - | BD |  | 25 |
| LIVE/DEAD | Efluor-506 | - | Invitrogen | 65-0866 | 0.5 |
| CD5 | BUV395 | UCHT1 | BD | 563546 | 1 |
| CD20 | BUV496 | 2H7 | BD | 749954 | 5 |
| IgD | BUV563 | IA6-2 | BD | 741394 | 1 |
| CD138 | BUV661 | MI15 | BD | 749873 | 1 |
| IgM | BUV737 | UCH-B1 | Bd | 748928 | 0.5 |
| CD14 | BUV805 | M5E2 | BD | 612902 | 3 |
| IgG | BV421 | G18-145 | BD | 562581 | 2 |
| CD3 | BV510 | UCHT1 | BD | 566105 | 0.5 |
| CD56 | BV480 | NCAM16.2 | BD | 566124 | 0.5 |
| CXCR5** | BV605 | J252D4 | Biolegend | 356929 | 4 |
| CD19 | BV650 | SJ25C1 | Biolegend | 363028 | 1 |
| t-bet*** | BV711 | O4-46 | BD | 563320 | 5 |
| CD21 | BV786 | B-LY4 | BD | 740969 | 0.5 |
| RBD probe | AF488 |  | *In house* | NA | 0.4 |
| CD11c | BB700 | SHCL-3 | BD | 746106 | 1 |
| CD38 | BB790 | HIT2 | BD | *custom* | 3 |
| RBD probe | AF594 |  | *In house* | NA | 0.4 |
| Ki67*** | PE-Cy7 | B56 | BD | 561283 | 5 |
| IgA | APC Vio770 | IS11-8E10 | Miltenyi | 130-113-999 | 2.5 |
| CD27 | APC-R700 | M-T271 | BD | 566116 | 0.5 |
| S100A8/9*** | eF660 | CF-145 | Ebioscience | 50-9745-42 | 5 |
| * one test : 5M PBMC in 100µL staining buffer | | |  |  |  |
| ** added in cull culture (0.5ml) 15 min prior to cell collection for staining.  ***intranuclear or intracellular staining | | | | | |

**Supplementary Table 5: Flow cytometry panel used to detect AIM^+^ Spike-specific T cells**

| Antigen/Reagent | Fluorochrome | Clone | Manufacturer | Cat # | Volume/test (µL)* |
| --- | --- | --- | --- | --- | --- |
| Brilliant Stain buffer | - | - | BD | 563794 | 25 |
| LIVE/DEAD | Efluor-506 | - | Invitrogen | 65-0866 | 0.5 |
| CD40 block** | - | HB14 | Miltenyi | 130-094-133 | 1 |
| CD3 | BUV395 | UCHT1 | Bd | 563546 | 3 |
| CD4 | BUV496 | SK3 | BD | 612936 | 4 |
| CD27 | BUV661 | L128 | BD | 750167 | 0.5 |
| CCR6** | BUV737 | 11A9 | BD | 564377 | 0.5 |
| CXCR6** | BUV805 | 13B 1E5 | BD | 748448 | 2 |
| CXCR5** | BV421 | J252D4 | Biolegend | 356920 | 3 |
| CD14 | BV480 | M5E2 | BD | 746304 | 3 |
| CD19 | BV480 | HIB19 | BD | 746457 | 3 |
| CD16 | BV480 | 3G8 | BD | 566108 | 1 |
| CD20 | BV480 | 2G7 | BD | 566181 | 0.5 |
| CD56 | BV480 | NCAM16.2 | BD | 566124 | 0.5 |
| CD8 | BV570 | RPA-T8 | Biolegend | 301037 | 1 |
| CXCR3** | BV605 | G025H7 | Biolegend | 353728 | 0.5 |
| CD69 | BV650 | FN50 | Biolegend | 310934 | 2 |
| PD-1 | BV711 | EH12.2H7 | Biolegend | 329928 | 4 |
| HLA-DR | FITC | LN3 | Biolegend | 327005 | 0.5 |
| CD45RA | PerCP Cy5.5 | HI100 | BD | 563429 | 0.5 |
| CD38 | BB790 | HIT2 | BD | *custom* | 3 |
| CD40L | PE | TRAP1 | BD | 555700 | 5 |
| 41BB | PE-Dazzle594 | 4B4-1 | Biolegend | 309826 | 2 |
| CCR7** | PE-Cy7 | 3D12 | BD | 560922 | 1 |
| OX40 | APC | ACT35 | BD | 563473 | 2 |
| * one test : 1.7M PBMC in 100µL staining buffer or 170µL of culture media | | | | | |
| ** added in cull culture (0.5mL) 15 min prior to stimulation | | | | | |

**Supplementary Table 6. Flow cytometry antibody staining panel for intracellular cytokine detection**

| Target | Fluorochrome | Clone | Manufacturer | Detection | Catalog number | Volume per test (µL)* |
| --- | --- | --- | --- | --- | --- | --- |
| CD3 | BUV395 | UCHT1 | BD Biosciences | Surface | 563546 | 3 |
| CD4 | BUV496 | SK3 | BD Biosciences | Surface | 564651 | 4 |
| CD8 | BV570 | RPA-T8 | Biolegend | Surface | 344732 | 1 |
| CD14 | BUV805 | M5E2 | BD Biosciences | Surface | 612902 | 3 |
| CD16 | BV650 | 3G8 | Biolegend | Surface | 302042 | 3.5 |
| CD19 | APC-eFluor780 | HIB19 | ThermoFisher | Surface | 47-0199-42 | 0.5 |
| CD56 | BUV737 | NCAM16.2 | BD Biosciences | Surface | 564448 | 2.5 |
| CD69 | PerCP-eFluor710 | FN50 | ThermoFisher | Intracellular | 46-0699-42 | 4 |
| CD107a** | BV785 | H4A3 | Biolegend | During stimulation | 328644 | 1 |
| CD154**  (CD40L) | BV421 | TRAP1 | BD Biosciences | Intracellular | 563886 | 5 |
| Granzyme B** | AF700 | GB11 | BD Biosciences | Intracellular | 561016 | 1 |
| IFN-γ** | PE-Cy7 | B27 | BD Biosciences | Intracellular | 557643 | 4 |
| IL-2** | PE-Dazzle594 | MQ1-17H12 | Biolegend | Intracellular | 500344 | 3.5 |
| IL-10** | PE | JES3-9D7 | BD Biosciences | Intracellular | 554498 | 5 |
| IL-17A** | eFluor660 | eBio64CAP17 | ThermoFisher | Intracellular | 50-7178-42 | 5 |
| TNF-α** | AF488 | MAb11 | Biolegend | Intracellular | 502915 | 2 |

*One test: 2M PBMCs in 100µl staining buffer for surface and intracellular detection or 2M PBMCs in 500µl of media for staining during culture

**intranuclear or intracellular staining
